# Physical activity, low-grade inflammation, and psychological responses to the COVID-19 pandemic among older adults in England

**DOI:** 10.1101/2024.04.14.24305797

**Authors:** Martin N. Danka, Andrew Steptoe, Eleonora Iob

**Affiliations:** Centre for Longitudinal Studies, University College London, UK; Department of Behavioural Science and Health, University College London, UK

## Abstract

Mental health responses to the COVID-19 pandemic have been widely studied, but less is known about the potentially protective role of physical activity (PA) and the impact of low-grade inflammation. Using a sample of older adults from England, this study tested (1) if pre-pandemic PA and its changes during the pandemic were associated with mental health responses; (2) if older adults with low-grade inflammation experienced greater increases in depression and anxiety, compared to pre-pandemic levels; (3) if PA attenuated the association between inflammation and depression/anxiety. The study used data from the English Longitudinal Study of Ageing, a cohort study following a national sample aged 50+. Information on mental health and PA were collected before the pandemic (2016/17 and 2018/19) and during November and December 2020. Inflammation was ascertained using pre-pandemic C-reactive protein (CRP). Analyses were adjusted for sociodemographic and health-related factors and pre-pandemic mental health. Increasing PA from before to during the pandemic was linked to reduced odds of depression (*OR* = 0.955, 95%*CI* [0.937, 0.974]) and anxiety (*OR* = 0.954, 95%*CI* [0.927;0.982]). Higher pre-pandemic PA was associated with reduced odds of depression (*OR* = 0.964, 95%*CI* [0.948, 0.981]) and anxiety (*OR* = 0.976, 95%*CI* [0.953, 1.000]), whereas elevated CRP was associated with 1.343 times higher odds of depression (95%*CI* [1.100, 1.641]). PA did not attenuate the inflammation-depression association. The findings suggest that PA may contribute to psychological resilience among older adults, independently of inflammation. Further research is needed to explore the psychobiological pathways underlying this protective mechanism.

## INTRODUCTION

In 2019, depressive and anxiety disorders ranked among the top 25 leading causes of global and UK disease burden [1, 2]. The COVID-19 pandemic has further aggravated the situation, resulting in a global increase in the prevalence of depressive and anxiety disorders by 27.6% and 25.6% in 2020, respectively [3]. In the UK, longitudinal surveys showed a similar pattern, with psychological distress being highest during the initial stages of the pandemic and changing in relation to social policy, confidence in healthcare services, COVID-19 related stress, and perceived social support or loneliness [4, 5].

Older adults are vulnerable to experiencing negative impacts of public health crises such as the COVID-19 pandemic in multiple domains, including higher risk of mortality, reduced access to healthcare, social isolation, and financial struggles [6]. A previous analysis of the English Longitudinal Study of Ageing used data collected by both telephone interviews and online surveys and found a substantial increase in the prevalence of depressive and anxiety disorders in March-May 2020 compared to pre-pandemic levels, worsening further at time of the second national lockdown in November-December 2020 [7]. Considering the disproportionate impacts of the COVID-19 pandemic on older adults, it is a research priority to identify risk and protective factors to their mental health [8].

Physical activity (PA), defined as *“any bodily movement produced by skeletal muscles that results in energy expenditure”* [9], has been a recognised protective factor against depressive [10] and anxiety [11] disorders. Pandemic-related restrictions have been linked to decreases in PA and increases in sedentary behaviours in the general as well as older population [12, 13]. An analysis of the UK PROTECT cohort found that older adults who reported reductions in their PA compared to pre-pandemic levels were at an increased risk of depression and anxiety [14]. However, similarly to internet surveys on mental health, these studies collected online data and lacked pre-pandemic measures of PA. Therefore, it remains unclear whether PA may promote resilience to mental health issues among older adults in response to the COVID-19 pandemic.

Older age and PA might be linked to mental health through inflammatory pathways. The process of ageing is accompanied by chronic subclinical increases in inflammatory biomarkers (low-grade inflammation, LGI), which may in turn contribute to various physical as well as mental health issues [15]. Conversely, aerobic exercise interventions can reduce inflammatory biomarkers, including C-reactive protein (CRP), tumour necrosis factor alpha (TNF-α), and interleukin-6 (IL-6) [16]. The role of inflammation in mental health has been corroborated by both animal and human studies, with growing evidence that inflammation may be involved in the pathogenesis of depression [17]. Although inflammation has been proposed to play a role in anxiety as well [18, 19], the evidence is more limited [20–26].

Unlike experimentally triggered acute inflammation, LGI can be less intense and prolonged. Chronically elevated inflammatory biomarkers might increase the susceptibility to mental health disorders by inducing structural and functional changes in affective brain circuits [27]. LGI could prime individuals to various mental health responses, which would manifest under challenging or stressful circumstances. In that case, LGI would be an especially relevant risk factor when encountering novel and unpredictable stressors, including those that have emerged during the COVID-19 pandemic. A previous study has found support for this mechanism, showing that older adults with elevated inflammation before the pandemic were more likely to develop depression [28]. Under these circumstances, the inflammation-anxiety link might also become more apparent, although this relationship remains to be tested.

Although the beneficial effects of PA on mental health could be partially explained by the reduction in inflammatory biomarkers, direct evidence is missing. In a longitudinal study of older adults, inflammation accounted for only a small portion of the PA effects on depressive symptoms [29]. A randomised-controlled trial of patients with major depressive disorder found greater improvements in depression symptoms following exercise intervention in those with higher IL-6 at baseline, whilst no changes were found in the inflammatory biomarkers before and after the intervention [30]. Notably, as PA operates through multiple pathways, the anti-inflammatory mechanisms might only be relevant to mental health if inflammation is present in the first place. Additionally, other mechanisms of PA might compensate for the negative effects of inflammation on the brain, such as the upregulation and altered signalling of the brain-derived neurotrophic factor, a protein involved in neurogenesis and synaptic stability [31, 32]. Therefore, PA could act as an effect modifier, buffering the association between LGI and mental health. However, studies directly testing the modifying role of PA are missing.

To address these evidence gaps, we aimed to test three main hypotheses using observational data from a sample of older adults living in England. Firstly, we postulated that higher pre-pandemic PA and increases in PA from before to during the pandemic would be associated with reduced mental health responses to the pandemic (depression and anxiety). Second, we hypothesized that higher pre-pandemic LGI would be associated with an increased risk of depression and anxiety during the pandemic. Third, we expected that higher levels of pre-pandemic PA and increases in PA from before to during the pandemic would attenuate the association between LGI and depression/anxiety.

## MATERIAL AND METHODS

### Design

This study used data from the English Longitudinal Study of Ageing (ELSA), a prospective multidisciplinary cohort study following a nationally representative sample of adults aged 50 years and above living in England. Data on socioeconomic and health variables are obtained biennially via computer-assisted face-to-face interviews and self-completion questionnaires. Every four years, biomedical data are collected during nurse visits. In 2020, the COVID-19 sub-study was launched to monitor the impacts of the COVID-19 pandemic on older population of England, collecting data by telephone or online self-completion interviews. Details on sampling and recruitment for all ELSA waves can be found on the ELSA website (https://www.elsa-project.ac.uk/). All participants provided informed consent. The data from ELSA waves can be accessed through the UK Data Service. Ethical approval was obtained from the National Research Ethics Service for the main ELSA waves and the University College London Research Ethics Committee for the COVID-19 sub-study.

In the current study, we derived a dataset comprising participants who completed their final nurse visit prior to the pandemic. This encompassed either Wave 8 (May 2016 – June 2017) or Wave 9 (June 2018 – July 2019), as data collection was divided between these two waves due to financial constraints. Participants were excluded if they had blood clotting disorders or history of fits or convulsions, as they were ineligible to provide a blood sample (532 participants). We also excluded individuals with inflammatory biomarker levels indicating an acute infection or pathology (CRP ≥10 mg/L, 234 participants). The final analytical sample consisted of 5 829 individuals (Supplementary Fig. 1). Pandemic mental health outcomes and PA variables were extracted from Wave 2 of the COVID-19 sub-study (November-December 2020), as the same PA measures were also collected at Wave 8 and Wave 9, allowing to compare PA changes from before to during the pandemic. Some confounders were sourced from the last wave before the exposure, either Wave 7 or Wave 8, to avoid overadjustment bias [33] The timing of all variables is shown in Supplementary Figure 2. The study protocol was pre-registered prior to analysing the data with the Open Science Framework registry [34]. Modifications to the analyses are detailed in Supplementary Table 1. We followed the STROBE guidelines to enhance the reporting of our study [35].

### Measures

#### Pandemic mental health (outcome)

Depression was measured using the 8-item version of the Center for Epidemiologic Studies-Depression scale (CES-D 8). This scale collects information on eight symptoms of depression (e.g., feeling lonely, feeling depressed, experiencing restless sleep) and their frequency in the past week. The full 20-item version of CES-D is a well-established tool for screening depression in the general population [36]. The 8-item version has been validated in populations of older adults [37, 38]. To assess clinically significant depressive symptoms indicating the presence of depressive disorders, we used a cut-off score of four or more symptoms that is equivalent to the clinical cut-off score of 16 on the 20-item scale [39].

Anxiety was ascertained using the 7-item Generalized Anxiety Disorder (GAD-7) scale. The GAD-7 collects information on anxiety symptoms in the past two weeks and their frequency (such as feeling nervous, anxious, or on edge, not being able to stop worrying). The scale is a gold standard for screening anxiety disorders in adult populations, with a cut-off score of 10 or more for identifying clinically significant symptoms of anxiety [40]. This cut-off score was used to re-code GAD-7 scores into a binary variable.

#### Physical activity (exposure)

Participants were asked how often they engaged in mildly energetic, moderately energetic, and vigorous PA. Prompt cards were used to illustrate examples of each PA category (e.g., laundry for mild, gardening for moderate, and swimming for vigorous PA). The response options included *hardly ever, or never (0), one to three times a month (1), once a week (2), more than once a week (3).* To capture the overall PA engagement of a person, we computed a continuous PA index as the sum of the responses, adding higher weights to more intense PA engagement (PA_index_ = PA_mild_ + 2 × PA_moderate_ + 3 × PA_vigorous_). The effects of PA changes were derived statistically, using adjustment for past exposure levels (PA_index_ at W2 of the COVID-19 sub-study adjusted for PA_index_ at baseline). This approach, comparable to using change scores adjusted for baseline values, yields interpretable effects of change-in-exposure as recently defined within the potential outcomes framework [41].

#### Low-grade inflammation (exposure)

High-sensitivity plasma CRP was used as a biomarker of inflammation. During the nurse visit, at least three small-sized tubes (2-6 mL each) of blood were obtained from each participant. The samples were stored and analysed at the Royal Victoria Infirmary laboratory using N Latex CRP highly sensitive mono immunoassay (Behring Nephelometer II Analyser). We used the cut-off score of CRP ≥3 mg/L to indicate clinically significant subclinical systemic inflammation (referred to as LGI in this paper). We used this threshold because higher CRP concentrations are associated with adverse health outcomes, such as cardiovascular disease. In contrast, values below this threshold might not be consistently linked to clinical outcomes, as they could represent physiological variations within the normal range [42].

#### Confounders

The analyses were adjusted for pre-pandemic mental health to isolate the mental health responses to the pandemic. The same measure (CES-D 8) was used for pre-pandemic depression. GAD-7 was only added in the COVID-19 sub-study. Therefore, to account for pre-pandemic differences in anxiety, we used the 11-point (0-10) anxiety item of the Office for National Statistics well-being measure (ONS-4), which evaluates how anxious participants felt the previous day on a scale from 0 (“not at all”) to 10 (“completely”). As this scale does not have a comparable cut-off score to the GAD-7, we used the ONS anxiety tool as a continuous variable. Details on the ONS-4 measure can be found on the Government Statistical Service website: https://gss.civilservice.gov.uk/policy-store/personal-well-being/#dissemination-output-

As confounders, we included sociodemographic variables (age, sex, ethnicity, education, partnership status, household wealth) and health-related factors (having reported a longstanding illness that limits daily activities, smoking status, alcohol consumption). Self-rated weight at baseline was used as a confounder in a sensitivity analysis. Because baseline PA and LGI occurred before the pandemic, no events or conditions arising during the pandemic, including SARS-CoV-2 infection, could act as confounders in their relationship with mental health. Therefore, pandemic influences were not included as covariates to avoid overadjustment and collider bias [43]. A more detailed overview of all confounders is provided in Supplementary Table 2.

### Analyses

#### Main analyses

A series of logistic regression models were used to test the research hypotheses. Separate models were tested for clinically significant depressive and anxiety symptoms measured during the pandemic as the outcome variables. The models accounted for pre-pandemic depression or anxiety and the outlined confounders. Five models were fitted to each outcome variable, incorporating different exposure and/or effect modifier variables: LGI (Model 1), pre-pandemic PA score (Model 2), LGI, pre-pandemic PA score, and their product term (Model 3), PA changes (isolated as pandemic PA adjusted for pre-pandemic PA as a covariate, Model 4), and PA changes, LGI, and a product term between pandemic PA and LGI (Model 5). A wave indicator was added to account for different exposure and confounder timing among the participants.

Assumptions were checked with diagnostic plots of scaled residuals using the DHARMa package v0.4.5 in R [44].

Each model provided odds ratios (*OR*s), their 95% confidence intervals, and *p*-values using Wald test. To visualise the results and supplement the interpretation of interactions, we plotted the average marginal effects at counterfactual exposure values (predicted probabilities marginalised over the distribution of confounders) together with their 95% confidence intervals using the avg_predictions command from the marginaleffects package v0.18.0 [45, 46]. As the outcomes had a high prevalence (>10%), we also assessed if *OR*s yielded similar results to risk ratios (*RR*s). *RR*s were approximated using modified Poisson regression with sandwich estimator for variance [47].

#### Missing data

Missing data on the exposures, covariates, and outcomes, including missingness in the outcome due to attrition, were handled using multivariate imputation by chained equations (MICE) with the *mice* package v3.16.0 [48]. We used 30 imputation datasets and 30 iterations. The estimates were pooled according to Rubin’s rules [49]. Details can be found in Supplement 1 and the pre-registration protocol [34].

#### Inference criteria

We followed guidelines aiming to improve statistical inference [50, 51]. *P*-values were reported as continuous quantities. Whilst we did consider the conventional alpha level of .05, we deliberately avoided the term ‘statistically significant’ and used additional metrics to supplement the interpretation of our findings, namely *s-*values, *OR*s and their 95% confidence intervals, and minimum detectable effects (*MDE*s) from sensitivity power analyses. The s-value is a re-expression of the p-value as the equivalent number of heads in a fair coin toss, recommended for providing a more intuitive scaling [52]. *MDE* represents the smallest effect (in this case, *OR_MDE_*) that could be detected with given power and sample size [53]. Confidence intervals were interpreted as compatibility intervals, meaning all the included effects were seen as highly compatible with the data [54]. For further details on these metrics, see Supplement 1.

#### Sensitivity analyses

We conducted six sensitivity analyses to check the robustness of our findings. Firstly, we re-ran the main models for complete cases to assess if the results are consistent with those of imputed datasets. Secondly, we re-ran the models with PA re-coded into a binary variable indicating if a participant engaged in moderate or vigorous PA at least once a week (high/low PA). To assess PA changes, we combined the two binary variables (before and during the pandemic), resulting in four binary variables (low-low, low-high, high-low, high-high). These variables were used by Hamer et al. [55] who found that high PA and changes in PA from low to high over a four-year period were associated with indicators of healthy ageing independently of sociodemographic and lifestyle factors. Thirdly, we repeated the main analyses using linear regression with a sandwich variance estimator using the CES-D 8 and GAD-7 scores as continuous outcomes. Fourth, we adjusted the main models for self-reported weight at baseline. We did not include this confounder in the main analyses, as we suspected it may lie on the causal pathway between inflammation or PA and mental health, and therefore could introduce overadjustment bias [56]. Fifth, to assess the effects of PA immediately before the pandemic, we re-ran the analyses for the main effects of PA using Wave 9 measures from everyone who attended COVID-19 Wave 2 of ELSA, with no other exclusions applied. This allowed us to use existing longitudinal survey weights to generalise the findings to the population of interest and address attrition, limiting imputations to item-missing data only. Lastly, we conducted the main analysis in full adherence to the pre-registration protocol to maintain transparency.

## RESULTS

### Descriptive statistics

Descriptive statistics are reported in Table 1. Most participants were white, partnered, and educated at a secondary level. Average age was 67.9 years (*SD* = 9.9) and 57.0% of the participants were female. 30.2% of the participants had at least one health condition limiting their daily activities. LGI was common, with approximately 22.5% participants showing CRP levels elevated above the cut-off score. At the time of pre-pandemic assessment, most participants frequently engaged in mild and moderate PA, with 81.7% and 65.6% reporting to have engaged ‘more than once a week’, respectively. By November-December 2020, PA levels had decreased, with changes being most apparent in mild-intensity PA engagement (Supplementary Figure 3). When using the derived PA index score, PA levels decreased, on average, by 0.8 points, but the changes varied considerably across participants (*SD* = 5.2).

**Table 1:** Descriptive statistics.

| Characteristic | % (count) | Mean (SD) | Median (IQR)<sup>†</sup> | Missing % (count) |
| --- | --- | --- | --- | --- |
| **Wave at baseline** |  |  |  |  |
| Wave 8 | 53.1 (3 095) | - | - | None |
| Wave 9 | 46.9 (2 734) | - | - | None |
| **Sociodemographic characteristics** |  |  |  |  |
| Sex: Female | 57.0 (3 320) | - | - | None |
| Age (years) | - | 67.9 (9.9) | - | None |
| Ethnicity: White | 96.0 (1 516) | - | - | <0.001 (2) |
| Partnership: Partnered | 68.0 (3 960) | - | - | <0.001 (2) |
| Education |  |  |  | 8.8 (512) |
| *Less than upper secondary* | 22.8 (1 213) | - | - | - |
| *Upper secondary and vocational training* | 55.0 (2 926) | - | - | - |
| *Tertiary* | 22.2 (1 178) | - | - | - |
| Wealth tertiles |  |  |  | 6.3 (370) |
| *First* | 33.3 (1 816) | - | - | - |
| *Second* | 34.4 (2 926) | - | - | - |
| *Third* | 32.3 (1,178) | - | - | - |
| **Health-related factors** |  |  |  |  |
| Limiting longstanding illness | 30.2 (1 516) | - | - | 13.8 (802) |
| Smoker | 9.3 (465) | - | - | 13.8 (806) |
| Alcohol consumption |  |  |  | 21.2 (1 235) |
| *Three or more times a week* | 34.3 (1 576) | - | - | - |
| *Once or twice a week* | 24.9 (1 146) | - | - | - |
| *Less than once a week or not at all in a year* | 40.7 (1 872) | - | - | - |
| **Exposures** |  |  |  |  |
| Low-grade inflammation (*hsCRP* $\geq 3$ mg/L) | 22.5 (1 080) | - | - | 17.7 (1 031) |
| *hsCRP (mg/L)* | - | 2.0 (2.0) | 1.2 (2.1) | 17.7 (1 031) |
| Physical activity (PA) |  |  |  |  |
| *PA index (pre-pandemic)* | - | 10.1 (5.5) | - | <0.001 (1) |
| *PA index (pandemic)* | - | 10.0 (5.4) | - | 26.2 (1 525) |
| *PA index difference* | - | -0.8 (5.2) | - | 26.2 (1 525) |
| **Mental health outcomes** |  |  |  |  |
| Depressive symptoms (pre-pandemic) |  |  |  |  |
| *Elevated (CES-D 8 $\geq 4$)* | 11.4 (659) | - | - | 6.6 (57) |
| *CES-D 8* | - | 1.3 (1.8) | 1.0 (2.0) | 6.6 (57) |
| Depressive symptoms (pandemic) |  |  |  |  |
| *Elevated (CES-D 8 $\geq 4$)* | 23.9 (1 014) | - | - | 27.2 (1 583) |
| *CES-D 8* | - | 2.1 (2.3) | 1.0 (3.0) | 27.2 (1 583) |
| Anxiety symptoms (pre-pandemic) |  |  |  |  |
| *ONS-4* | - | 2.5 (2.6) | 2.0 (4.0) | 10.3 (1 288) |
| Anxiety symptoms (pandemic) |  |  |  |  |
| *Elevated (GAD-7 $\geq 10$)* | 8.8 (372) | - | - | 27.2 (1 587) |
| *GAD-7* | - | 3.2 (4.2) | 2.0 (5.0) | 27.2 (1 587) |
| **Additional confounders** |  |  |  |  |
| Self-rated weight |  |  |  | 0.5 (17) |
| *Too light* | 3.2 (222) | - | - | - |
| *About the right weight* | 44.4 (2 556) | - | - | - |
| *Too heavy* | 52.4 (3 034) | - | - | - |

Note. Descriptive statistics computed from complete-case data. *IQR* = interquartile range; *SD* = standard deviation.
† Listed for continuous variables with skewness or kurtosis $\geq 1.0$

The proportion of participants with clinically significant depressive symptoms increased from before to during the pandemic by 12.5%. We could not assess changes in proportions of participants with clinically significant anxiety symptoms, as ONS-4 and GAD-7 may not offer comparable cut-offs. During the pandemic, a larger proportion of participants experienced clinically significant depressive symptoms (23.9%) compared to anxiety symptoms (8.8%). Descriptive statistics computed across the imputed datasets were comparable to those obtained from the complete-case data (Supplementary Table 3). Descriptive statistics for the full Wave 8 and Wave 9 samples were also similar to those obtained from the analytical sample, although their prevalence of clinically significant depressive symptoms at baseline was higher (Supplementary Table 4).

### PA, LGI, and depressive symptoms

The full models for depression are reported in Table 2 and plotted in Fig. 1. Unadjusted associations are provided in the supplements (Supplementary Table 5). Participants with LGI before the pandemic had 1.34 times increased odds of developing depressive disorders during the pandemic independently of pre-pandemic depression and confounders (95% CI [1.100; 1.641], *p* = 0.004, *s* = 7.92). The adjusted odds of developing depression decreased with higher pre-pandemic PA engagement (*OR* = 0.964, 95% CI [0.948; 0.981], *p* = <0.001, *s* = 14.98); a 6- point increase in the PA index (equivalent to engaging more than once a week in moderately energetic or at least once a week in vigorous PA) corresponded to a 20% reduction in the adjusted odds of developing depression. Similarly, increasing PA levels from before to during the pandemic by 6 points corresponded to a 24% decrease in the adjusted odds of having clinically significant depressive symptoms during the pandemic, although the evidence was weaker (*OR* = 0.955, 95% CI [0.937, 0.974], *p* <0.001, *s* = 17.50). The models found little support for pre-pandemic PA modifying the association between inflammation and depression (*OR*_PA * LGI_ = 0.993, 95% CI [0.959; 1.029], *p* = 0.710, *s* = 0.49). Similarly, the point estimate and confidence interval suggested no effect modification by PA changes (*OR*_PAchange * LGI_ = 0.992, 95% CI [0.959; 1.025], *p* = 0.619, *s* = 0.693).

**Fig. 1:**
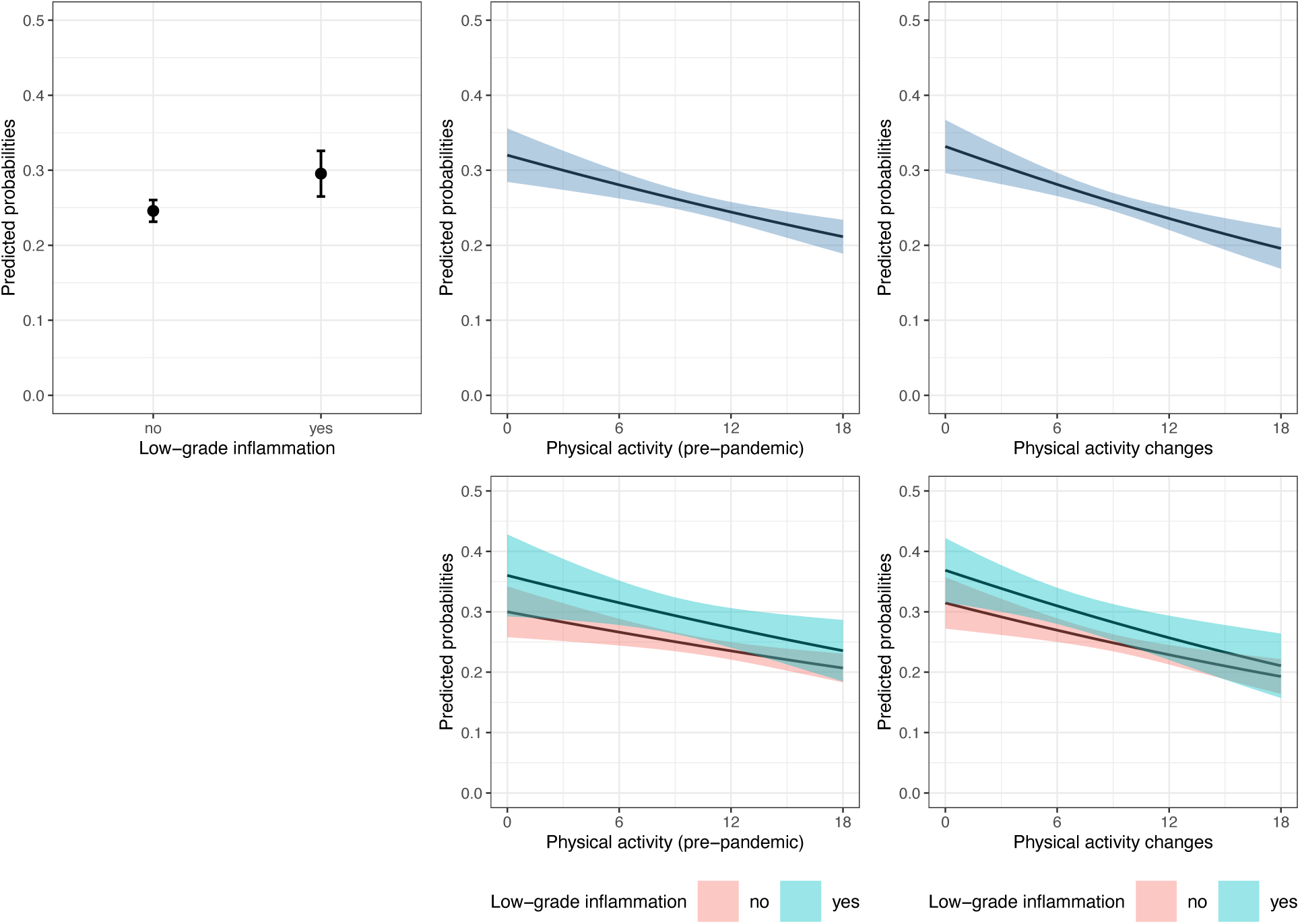
Plots of average marginal probabilities of depression from adjusted logistic regression models. *Note.* The plots show average marginal probabilities at counterfactual values (predicted probabilities marginalised over the distribution of confounders) and their 95% confidence intervals of experiencing clinically significant depressive symptoms during the pandemic (November-December 2020) depending on pre-pandemic low-grade inflammation (2016/17 or 2018/19), physical activity engagement (2016/17 or 2018/19), physical activity changes from before to during the pandemic, and interactions between physical activity and low-grade inflammation. The predictions were pooled from logistic regression models fitted to 30 imputed datasets (sample N = 5,829) and adjusted for pre-pandemic depressive symptoms, sex, age, ethnicity, education, partnership status, household wealth, having a limiting longstanding illness, smoking status, and alcohol consumption

**Table 2:** Results of the main adjusted logistic regression models.

| Variable | OR [95% CI] | SE | p | s | OR<sub>MDE</sub> | RR [95% CI] |
| --- | --- | --- | --- | --- | --- | --- |
| **Outcome: Depression** |  |  |  |  |  |  |
| Model 1 |  |  |  |  |  |  |
| LGI | 1.343 [1.100; 1.641] | 0.10 | 0.004 | 7.92 | 1.198 | 1.200 [1.066; 1.350] |
| Model 2 |  |  |  |  |  |  |
| PA | 0.964 [0.948; 0.981] | 0.01 | <0.001 | 14.98 | 0.981 | 0.977 [0.966; 0.988] |
| Model 3 |  |  |  |  |  |  |
| LGI | 1.367 [0.911; 2.052] | 0.21 | 0.130 | 2.95 | - | 1.147 [0.934; 1.408] |
| PA | 0.969 [0.950; 0.988] | 0.01 | 0.001 | 9.47 | - | 0.978 [0.966; 0.990] |
| LGI * PA | 0.993 [0.959; 1.029] | 0.02 | 0.710 | 0.49 | - | 1.002 [0.983; 1.023] |
| Model 4 |  |  |  |  |  |  |
| PA change | 0.955 [0.937; 0.974] | 0.01 | <0.001 | 17.50 | 0.980 | 0.971 [0.959; 0.983] |
| Model 5 |  |  |  |  |  |  |
| LGI | 1.319 [0.955; 1.822] | 0.16 | 0.093 | 3.43 | - | 1.133 [0.957; 1.341] |
| PA change | 0.959 [0.938; 0.980] | 0.01 | <0.001 | 11.88 | - | 0.972 [0.958; 0.986] |
| LGI * PA change | 0.992 [0.959; 1.025] | 0.02 | 0.619 | 0.693 | - | 1.001 [0.981; 1.021] |
| **Outcome: Anxiety** |  |  |  |  |  |  |
| Model 1 |  |  |  |  |  |  |
| LGI | 0.904 [0.669; 1.222] | 0.15 | 0.508 | 0.98 | 1.270 | 0.924 [0.733; 1.166] |
| Model 2 |  |  |  |  |  |  |
| PA | 0.976 [0.953; 1.000] | 0.012 | 0.049 | 4.35 | 0.972 | 0.982 [0.964; 1.002] |
| Model 3 |  |  |  |  |  |  |
| LGI | 1.148 [0.660; 1.995] | 0.28 | 0.622 | 0.69 | - | 1.096 [0.742; 1.620] |
| PA | 0.984 [0.958; 1.010] | 0.01 | 0.218 | 2.20 | - | 0.988 [0.967; 1.009] |
| LGI * PA | 0.964 [0.911; 1.020] | 0.03 | 0.197 | 2.34 | - | 0.973 [0.932; 1.014] |
| Model 4 |  |  |  |  |  |  |
| PA change | 0.954 [0.927; 0.982] | 0.01 | 0.002 | 9.30 | 0.970 | 0.964 [0.942; 0.987] |
| Model 5 |  |  |  |  |  |  |
| LGI | 1.008 [0.606; 1.678] | 0.26 | 0.975 | 0.04 | - | 0.986 [0.683; 1.423] |
| PA change | 0.959 [0.929; 0.989] | 0.02 | 0.009 | 6.78 | - | 0.967 [0.943; 0.992] |
| LGI * PA change | 0.977 [0.927; 1.030] | 0.03 | 0.390 | 1.36 | - | 0.985 [0.947; 1.024] |

*Note.* The models show associations between clinically significant depressive or anxiety symptoms during the pandemic (November-December 2020) and pre-pandemic low-grade inflammation (2016/17 or 2018/2019; Model 1), pre-pandemic physical activity engagement (2016/17 or 2018/2019; Model 2), changes in physical activity from before to during the pandemic (Model 4), and interactions between inflammation and physical activity as indicated (Model 3 and 5). The results were pooled from 30 imputed datasets (sample N = 5,829). Odds ratios, standard errors, and p-values were obtained from logistic regression models. Risk ratios were approximated using modified Poisson regression with a sandwich variance estimator. The models were adjusted for pre-pandemic mental health (depression or anxiety depending on the outcome), sex, age, ethnicity, education, partnership status, household wealth, having a limiting longstanding illness, smoking status, and alcohol consumption. Effects of PA changes were estimated using coefficients of pandemic PA adjusted for pre-pandemic PA and the outlined confounders. CI = confidence interval; LGI = low-grade inflammation; OR = adjusted odds ratio; OR<sub>MDE</sub> = minimum detectable odds ratio; PA = physical activity; RR = risk ratio, s = Shannon information value (surprisal value); SE = standard error.

### PA, LGI, and anxiety symptoms

Anxiety models are reported in Table 2 and plotted in Fig. 2. In contrast with depression, we did not find evidence of the association between LGI and clinically elevated anxiety symptoms (*OR* = 0.904, 95% CI [0.669; 1.222], *p* = 0.508, *s* = 0.98). Considering the *OR_MDE_* = 1.270 (also *OR_MDE_* = 0.765 if the association was negative), the analysis may have been underpowered to detect substantially smaller, yet meaningful associations of LGI with anxiety. The association strength between pre-pandemic PA levels and clinically significant anxiety symptoms was weaker and less precise compared to that with depression, with a 6-point increase in pre- pandemic PA levels corresponding to a 14% decrease in the adjusted odds of developing clinically significant anxiety symptoms (*OR* = 0.976, 95% CI [0.953, 1.000], *p* = 0.049, *s* = 4.35). Increasing PA levels by 6 points was associated with a 25% decrease in the adjusted odds of anxiety (*OR* = 0.954, 95% CI [0.927, 0.982], *p* = 0.002, *s* = 9.30). As for depression, the product term coefficients including pre-pandemic PA or PA changes were almost null and imprecise.

**Fig. 2:**
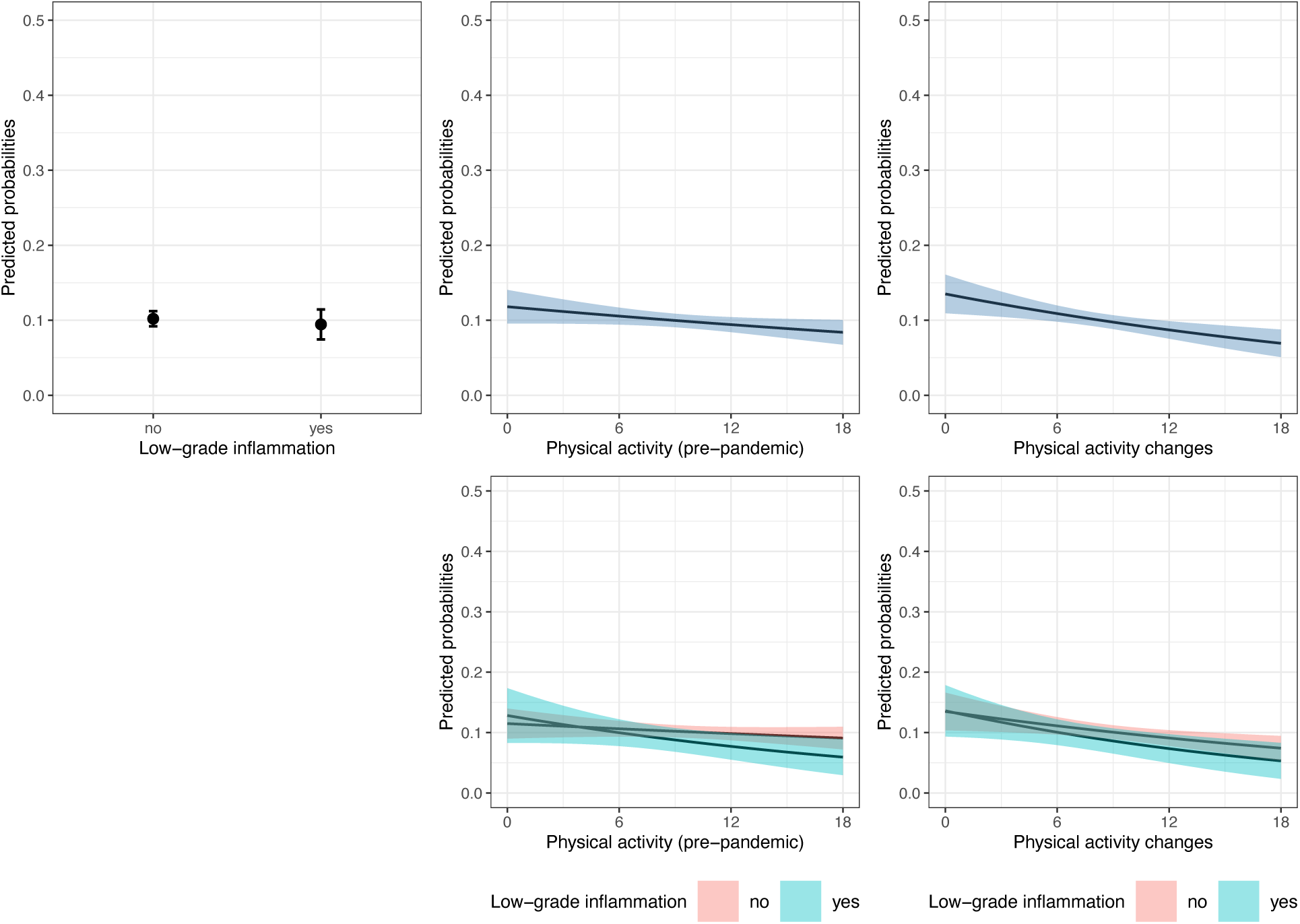
Plots of average marginal probabilities of anxiety from adjusted logistic regression models. *Note.* The plots show average marginal predicted probabilities at counterfactual values (predicted probabilities marginalised over the distribution of confounders) and their 95% confidence intervals of experiencing clinically significant anxiety symptoms during the pandemic (June to July 2020) depending on pre-pandemic low-grade inflammation (2016/17 or 2018/19), physical activity engagement (2016/17 or 2018/19), physical activity changes from before to during the pandemic, and interactions between physical activity and low-grade inflammation. The predictions were pooled from logistic regression models fitted to 30 imputed datasets (sample N = 5,829) and adjusted for pre-pandemic depressive symptoms, sex, age, ethnicity, education, partnership status, household wealth, having a limiting longstanding illness, smoking status, and alcohol consumption

### Sensitivity analyses

Across all models, the approximated *RR*s were comparable (although slightly smaller) to *ORs*.

The sensitivity analyses yielded results consistent with the main findings (Supplement 2).

## DISCUSSION

### Summary of findings

We found that higher pre-pandemic PA engagement was associated with reduced odds of developing clinically significant depressive and anxiety symptoms in response to the pandemic, irrespective of sociodemographic and health-related factors. Engagement in moderate-intensity PA more than once a week corresponded to an estimated 20% decrease in the adjusted risk of depression. A similar but less precise association was found for anxiety (14% decrease in the adjusted odds). Increases in PA were associated with reduction in the adjusted risk of depression and anxiety (24% and 25% decrease per moderate weekly PA engagement equivalent, respectively).

Older adults with elevated levels of CRP (LGI) before the pandemic had 1.34 times higher adjusted risk of developing clinically significant symptoms of depression and anxiety. The relationship between LGI and anxiety was smaller and imprecise. Higher pre-pandemic PA and increases in PA did not modify the LGI-depression/anxiety relationship, with estimated effects close to zero.

### Interpretation of the findings

Our findings align with evidence supporting the long-term protective role of PA on mental health. Although studies have established that decreases in PA during lockdowns are related to worsening of mental health [57], the overall impact of the COVID-19 pandemic becomes inevitably lost when no pre-pandemic measures are available. Our study addressed this gap, suggesting that PA may contribute to longer-term psychological resilience of older adults that persists in times of public health crises. We also found reductions in the risk of depressive and anxiety disorders for even small increases in PA. In our sensitivity analyses, we found similar associations when re-defining PA as a binary variable expressing moderate-to-vigorous PA engagement. This is in line with former research suggesting a dose-response relationship [58, 59]. However, a meta-analysis of eight randomised-controlled trials identified a non-linear association in studies focusing on adults aged 60+, suggesting no reduction in depressive symptoms with vigorous activity [60]. Whilst our diagnostic plots did not indicate a non-linear relationship, our sample included younger participants (50+) and we did not separately test the effects of vigorous PA.

In line with evidence for the role of inflammation in the pathogenesis of depression [17], our analysis revealed that pre-pandemic inflammation was associated with an increased risk of developing depressive symptoms during the pandemic, as evidenced by data from the second wave of the ELSA COVID-19 sub-study. Our findings align with those of Hamilton et al. [28], who identified a similar association using first-wave data from the ELSA COVID-19 sub-study, despite methodological differences between our studies. This consistency underscores the robustness of the link between inflammation and depression, but also demonstrates its persistence through the pandemic, making inflammation an important biological pathway connecting the process of ageing with vulnerability to depressive disorders in times of mandated social restrictions.

The relationship between inflammation and anxiety was weak and imprecise, so no robust conclusions can be drawn. The compatible effects were generally smaller than those found in depression, which aligns with previous studies. In an analysis of UK Biobank data, the association for the inflammation-anxiety relationship was weaker compared to that between inflammation and depression, and it was entirely attenuated following adjustment for depression. Conversely, the association between CRP and depression persisted even after the adjustment for anxiety, suggesting disorder specificity [26]. Considering that anxiety and depression disorders are highly comorbid [61, 62], inflammation could act through depression on anxiety.

To the best of our knowledge, this is the first study to have longitudinally tested the modifying role of PA on the association between LGI and mental health. This mechanism has previously been proposed due to interactions between physiological systems involved in exercise, inflammation, and depression, although its empirical support was limited [63]. PA has been implicated in increasing the expression of BDNF [31, 32], promoting the release of catecholamines [64], and modulating HPA axis hyperreactivity [65]. Conversely, inflammatory biomarkers can dysregulate these systems [66]. Contrary to this hypothesis, our findings do not support the notion of PA mitigating the LGI-depression link, suggesting PA and LGI might act independently of each other. In a former study, reduced inflammation explained only a small portion of the PA-mental health association [29]. An explanation could be that PA might reduce inflammation through mechanisms that simultaneously improve mental health outcomes. To illustrate, BDNF could exert both a neurotrophic as well as pro/anti-inflammatory activity [67]. Animal studies suggest that β-hydroxybutyrate, a ketone released during exercise, may also exert a simultaneous anti-inflammatory and anti-depressant activity [68–70].

Our findings could also reflect that plasma CRP levels may not directly mediate the neurobehavioral effects of systemic inflammation. Whilst plasma biomarkers of systemic inflammation are commonly used to examine associations between inflammation and mental health, their transport into the brain is limited by the blood-brain barrier (BBB), a semipermeable border lining the blood vessels of the brain that regulates the exchange of cells and molecules. Neuroinflammation can be promoted through several pathways, including the disruption of BBB, upregulation of cytokine transport across BBB, and signal transduction by barrier cells [71]. The interactions of CRP with BBB are not fully understood. Some evidence suggests that the association between CRP and depression could be confounded by other cytokines, such as IL- 6. In the systemic inflammatory response, IL-6 plasma levels increase prior to CRP. IL-6 then promotes CRP secretion into blood by the liver [72]. Unlike CRP, IL-6 has been documented to traverse BBB when elevated [73, 74]. In a Mendelian randomisation study, genetically predicted higher levels of IL-6 were related to increased depressive symptoms, whereas genetic instruments for elevated CRP showed protective effects in depression. This suggests that CRP may not directly increase the risk of depression. Instead, the associations found in observational studies could be explained by IL-6 and other cytokines acting on both CRP and depression [26].

### Strengths and limitations

Our study has several strengths. In our analysis, we used data from a nationally representative sample of adults aged 50 and above. Data collection was not limited to online interviews, so our sample did not exclude often-unrepresented older adults who do not use the internet [75]. Another strength is the use of validated scales to assess depressive and anxiety symptoms. Additionally, pre-pandemic data collection allowed us to estimate the overall increases in depressive and anxiety disorders in response to the pandemic and assess their longer-term associations with LGI and PA. We included a varied set of confounders and avoided overadjustment by extracting several variables from waves preceding the exposure. We also assessed the impacts of different modelling choices, demonstrating the robustness of most results.

Our findings should be interpreted in light of their limitations. First, PA and mental health variables were self-reported. Individuals often overestimate their PA levels [76], potentially leading to an underestimation of their association with mental health. While depression and anxiety were not verified through psychiatric interviews, we employed validated screening tools with established clinical cut-off scores. Second, the continuous PA index was derived using arbitrary weights because the PA measures in ELSA have not been validated or systematically combined. Nonetheless, employing a more conservative binary coding of PA levels produced similar results. Third, in our analysis of the GAD-7 outcome, we adjusted for pre-pandemic anxiety using a different measure, which may have led to reduced confounding control. Fourth, our study only used CRP as a biomarker of inflammation. As previously discussed, CRP may not fully capture the impact of systemic inflammation on the brain. Thus, the modifying role of PA on the inflammation-depression relationship might become more apparent when assessing other cytokines. Fifth, the measurement of pre-pandemic exposures varied, being taken at two distinct timepoints depending on the participant. Finally, selection bias could affect the study results because the participants who attended the nurse visit might not fully represent the target population, as suggested by higher baseline depression rates. To address these last two limitations in relation to PA and mental health associations, we conducted a sensitivity analysis using longitudinal survey weights with W9 data for PA. This analysis confirmed that the influence of pre-pandemic PA on mental health was robust to this form of selection bias.

### Suggestions for further research

Due to low precision of our estimates and inconsistent literature findings, future studies should re-examine the prospective associations between LGI and anxiety. Furthermore, to better understand the longer-term associations between PA and mental health among older adults, accelerometery or other objective PA measures would allow researchers to determine the recommended amount. As few studies tested the moderating role of PA on the relationship between LGI and mental health, replications are needed across populations and study designs. Future studies should implement other biomarkers of inflammation, such as IL-6 and TNF-α. Moreover, PA and inflammation interact through complex physiological pathways, rendering inferences from population-level data challenging. Researchers may wish to explore these relationships in greater depth, which may require novel approaches. To illustrate, new methods have been developed to integrate mediation and interaction [77].

## Conclusions

PA may contribute to psychological resilience of older adults in times of mandated social restrictions, making PA interventions a promising scalable approach to reducing the depression and anxiety burden associated with the COVID-19 pandemic. Intervention programmes aimed at improving the mental health of older populations may wish to promote different types of physical activities, such as walking, cycling, and hiking. During lockdowns, efforts should be made to maintain opportunities for PA engagement. Furthermore, inflammation is an age-related psychobiological mechanism underpinning vulnerability to depression in response to new environmental stressors, such as the COVID-19 pandemic. Whilst inflammation and PA may operate through shared physiological pathways on mental health, so far, studies have found no consistent pattern of interaction at population level. More longitudinal studies are needed to explore the interplay between PA, inflammation, and mental health.

## Data Availability

English Longitudinal Study of Ageing data are available for non-commercial research purposes via the UK Data Service: https://ukdataservice.ac.uk/

https://ukdataservice.ac.uk/

## Acknowledgments

Martin N. Danka is funded by the ESRC-BBSRC Soc-B Centre for Doctoral Training (ES/P000347/1). Eleonora Iob is supported by a Wellcome Trust Sir Henry Wellcome fellowship (222750/Z/21/Z, 2021–2025). The English Longitudinal Study of Ageing is funded by the Economic & Social Research Council (ESRC), the National Institute of Aging (R01AG017644), and UK government departments coordinated by the National Institute for Health Research (NIHR, Ref: 198-1074). The funders had no role in study design, data collection, data analysis, data interpretation, report writing, or manuscript submission.

## References

1. GBD 2019 Diseases and Injuries Collaborators. Global burden of 369 diseases and injuries in 204 countries and territories, 1990–2019: a systematic analysis for the Global Burden of Disease Study 2019. The Lancet. 2020;396:1204–1222.

2. McDaid D, Park A-L, Davidson G, John A, Knifton L, McDaid S, et al. The economic case for investing in the prevention of mental health conditions in the UK. Care Policy and Evaluation Centre, Department of Health Policy, London School of Economics and Political Science; 2022.

3. Santomauro DF, Mantilla Herrera AM, Shadid J, Zheng P, Ashbaugh C, Pigott DM, et al. Global prevalence and burden of depressive and anxiety disorders in 204 countries and territories in 2020 due to the COVID-19 pandemic. The Lancet. 2021;398:1700–1712.

4. Bu F, Steptoe A, Fancourt D. Depressive and anxiety symptoms in adults during the COVID- 19 pandemic in England: A panel data analysis over 2 years. PLOS Med. 2023;20:1–16.

5. Daly M, Robinson E. Psychological distress associated with the second COVID-19 wave: Prospective evidence from the UK Household Longitudinal Study. J Affect Disord. 2022;310:274–278.

6. Buffel T, Yarker S, Phillipson C, Lang L, Lewis C, Doran P, et al. Locked down by inequality: Older people and the COVID-19 pandemic. Urban Stud. 2021:004209802110410.

7. Zaninotto P, Iob E, Demakakos P, Steptoe A. Immediate and Longer-Term Changes in the Mental Health and Well-being of Older Adults in England During the COVID-19 Pandemic. JAMA Psychiatry. 2022;79:151.

8. Holmes EA, O’Connor RC, Perry VH, Tracey I, Wessely S, Arseneault L, et al. Multidisciplinary research priorities for the COVID-19 pandemic: a call for action for mental health science. Lancet Psychiatry. 2020;7:547–560.

9. Caspersen CJ, Powell KE, Christenson GM. Physical activity, exercise, and physical fitness: definitions and distinctions for health-related research. Public Health Rep. 1985;100:126– 131.

10. Schuch FB, Vancampfort D, Firth J, Rosenbaum S, Ward PB, Silva ES, et al. Physical Activity and Incident Depression: A Meta-Analysis of Prospective Cohort Studies. Am J Psychiatry. 2018;175:631–648.

11. McDowell CP, Dishman RK, Gordon BR, Herring MP. Physical Activity and Anxiety: A Systematic Review and Meta-analysis of Prospective Cohort Studies. Am J Prev Med. 2019;57:545–556.

12. Oliveira MR, Sudati IP, Konzen VDM, de Campos AC, Wibelinger LM, Correa C, et al. Covid-19 and the impact on the physical activity level of elderly people: A systematic review. Exp Gerontol. 2022;159:111675.

13. Stockwell S, Trott M, Tully M, Shin J, Barnett Y, Butler L, et al. Changes in physical activity and sedentary behaviours from before to during the COVID-19 pandemic lockdown: a systematic review. BMJ Open Sport Exerc Med. 2021;7:e000960.

14. Creese B, Khan Z, Henley W, O’Dwyer S, Corbett A, Vasconcelos Da Silva M, et al. Loneliness, physical activity, and mental health during COVID-19: a longitudinal analysis of depression and anxiety in adults over the age of 50 between 2015 and 2020. Int Psychogeriatr. 2021;33:505–514.

15. Franceschi C, Campisi J. Chronic Inflammation (Inflammaging) and Its Potential Contribution to Age-Associated Diseases. J Gerontol A Biol Sci Med Sci. 2014;69:S4–S9.

16. Zheng G, Qiu P, Xia R, Lin H, Ye B, Tao J, et al. Effect of Aerobic Exercise on Inflammatory Markers in Healthy Middle-Aged and Older Adults: A Systematic Review and Meta-Analysis of Randomized Controlled Trials. Front Aging Neurosci. 2019;11:98.

17. Miller AH, Raison CL. The role of inflammation in depression: from evolutionary imperative to modern treatment target. Nat Rev Immunol. 2016;16:22–34.

18. Michopoulos V, Powers A, Gillespie CF, Ressler KJ, Jovanovic T. Inflammation in Fear- and Anxiety-Based Disorders: PTSD, GAD, and Beyond. Neuropsychopharmacology. 2017;42:254–270.

19. Raison CL, Miller AH. Pathogen–Host Defense in the Evolution of Depression: Insights into Epidemiology, Genetics, Bioregional Differences and Female Preponderance. Neuropsychopharmacology. 2017;42:5–27.

20. Baune BT, Smith E, Reppermund S, Air T, Samaras K, Lux O, et al. Inflammatory biomarkers predict depressive, but not anxiety symptoms during aging: The prospective Sydney Memory and Aging Study. Psychoneuroendocrinology. 2012;37:1521–1530.

21. Copeland WE, Shanahan L, Worthman C, Angold A, Costello EJ. Generalized anxiety and C-reactive protein levels: a prospective, longitudinal analysis. Psychol Med. 2012;42:2641– 2650.

22. Costello H, Gould RL, Abrol E, Howard R. Systematic review and meta-analysis of the association between peripheral inflammatory cytokines and generalised anxiety disorder. BMJ Open. 2019;9:e027925.

23. Grigoleit J-S, Kullmann JS, Wolf OT, Hammes F, Wegner A, Jablonowski S, et al. Dose-Dependent Effects of Endotoxin on Neurobehavioral Functions in Humans. PLoS ONE. 2011;6:e28330.

24. Lasselin J, Elsenbruch S, Lekander M, Axelsson J, Karshikoff B, Grigoleit J-S, et al. Mood disturbance during experimental endotoxemia: Predictors of state anxiety as a psychological component of sickness behavior. Brain Behav Immun. 2016;57:30–37.

25. Reichenberg A, Yirmiya R, Schuld A, Kraus T, Haack M, Morag A, et al. Cytokine-Associated Emotional and Cognitive Disturbances in Humans. Arch Gen Psychiatry. 2001;58:445.

26. Ye Z, Kappelmann N, Moser S, Davey Smith G, Burgess S, Jones PB, et al. Role of inflammation in depression and anxiety: Tests for disorder specificity, linearity and potential causality of association in the UK Biobank. EClinicalMedicine. 2021;38:100992.

27. Won E, Kim Y-K. Neuroinflammation-Associated Alterations of the Brain as Potential Neural Biomarkers in Anxiety Disorders. Int J Mol Sci. 2020;21:6546.

28. Hamilton OS, Cadar D, Steptoe A. Systemic inflammation and emotional responses during the COVID-19 pandemic. Transl Psychiatry. 2021;11:626.

29. Hamer M, Molloy GJ, de Oliveira C, Demakakos P. Leisure time physical activity, risk of depressive symptoms, and inflammatory mediators: The English Longitudinal Study of Ageing. Psychoneuroendocrinology. 2009;34:1050–1055.

30. Rethorst CD, Toups MS, Greer TL, Nakonezny PA, Carmody TJ, Grannemann BD, et al. Pro-inflammatory cytokines as predictors of antidepressant effects of exercise in major depressive disorder. Mol Psychiatry. 2013;18:1119–1124.

31. Sleiman SF, Henry J, Al-Haddad R, El Hayek L, Abou Haidar E, Stringer T, et al. Exercise promotes the expression of brain derived neurotrophic factor (BDNF) through the action of the ketone body β-hydroxybutyrate. eLife. 2016;5:e15092.

32. Szuhany KL, Bugatti M, Otto MW. A meta-analytic review of the effects of exercise on brain-derived neurotrophic factor. J Psychiatr Res. 2015;60:56–64.

33. VanderWeele TJ. Principles of confounder selection. Eur J Epidemiol. 2019;34:211–219.

34. Danka MN, Iob E. Physical activity, low-grade inflammation, and psychological responses to the COVID-19 pandemic among older adults in England. 2022. 1 November 2022. 10.17605/OSF.IO/XJFYZ.

35. von Elm E, Altman DG, Egger M, Pocock SJ, Gøtzsche PC, Vandenbroucke JP. The Strengthening the Reporting of Observational Studies in Epidemiology (STROBE) statement: guidelines for reporting observational studies. J Clin Epidemiol. 2008;61:344– 349.

36. Radloff LS. The CES-D Scale: A Self-Report Depression Scale for Research in the General Population. Appl Psychol Meas. 1977;1:385–401.

37. Karim J, Weisz R, Bibi Z, ur Rehman S. Validation of the Eight-Item Center for Epidemiologic Studies Depression Scale (CES-D) Among Older Adults. Curr Psychol. 2015;34:681–692.

38. Missinne S, Vandeviver C, Van de Velde S, Bracke P. Measurement equivalence of the CES-D 8 depression-scale among the ageing population in eleven European countries. Soc Sci Res. 2014;46:38–47.

39. Steffick DE. Documentation of Affective Functioning Measures in the Health and Retirement Study. Institute for Social Research, University of Michigan; 2000.

40. Spitzer RL, Kroenke K, Williams JBW, Löwe B. A Brief Measure for Assessing Generalized Anxiety Disorder: The GAD-7. Arch Intern Med. 2006;166:1092.

41. Katsoulis M, Lai AG, Kipourou DK, Gomes M, Banerjee A, Denaxas S, et al. On the estimation of the effect of weight change on a health outcome using observational data, by utilising the target trial emulation framework. Int J Obes. 2023;47:1309–1317.

42. Pearson TA, Mensah GA, Alexander RW, Anderson JL, Cannon RO, Criqui M, et al. Markers of Inflammation and Cardiovascular Disease: Application to Clinical and Public Health Practice: A Statement for Healthcare Professionals From the Centers for Disease Control and Prevention and the American Heart Association. Circulation. 2003;107:499– 511.

43. Van Zwieten A, Tennant PWG, Kelly-Irving M, Blyth FM, Teixeira-Pinto A, Khalatbari-Soltani S. Avoiding overadjustment bias in social epidemiology through appropriate covariate selection: a primer. J Clin Epidemiol. 2022;149:127–136.

44. Hartig F. DHARMa: Residual Diagnostics for Hierarchical (Multi-Level / Mixed) Regression Models. 2022.

45. Arel-Bundock V. Marginaleffects: Predictions, Comparisons, Slopes, Marginal Means, and Hypothesis Tests. 2024.

46. Hernán M, Robins J. Causal Inference: What If. Boca Raton: Chapman & Hall/CRC; 2020.

47. Zou G. A Modified Poisson Regression Approach to Prospective Studies with Binary Data. Am J Epidemiol. 2004;159:702–706.

48. van Buuren S, Groothuis-Oudshoorn K. mice: Multivariate Imputation by Chained Equations in *R*. J Stat Softw. 2011;45.

49. Rubin DB. Multiple imputation for nonresponse in surveys. Hoboken, N.J: Wiley-Interscience; 2004.

50. Wasserstein RL, Lazar NA. The ASA Statement on p-Values: Context, Process, and Purpose. Am Stat. 2016;70:129–133.

51. Wasserstein RL, Schirm AL, Lazar NA. Moving to a World Beyond “ *p* &lt; 0.05”. Am Stat. 2019;73:1–19.

52. Greenland S. Valid *P* -Values Behave Exactly as They Should: Some Misleading Criticisms of *P* -Values and Their Resolution With *S* -Values. Am Stat. 2019;73:106–114.

53. Lakens D. Sample Size Justification. Collabra Psychol. 2022;8:33267.

54. Amrhein V, Trafimow D, Greenland S. Inferential Statistics as Descriptive Statistics: There Is No Replication Crisis if We Don’t Expect Replication. Am Stat. 2019;73:262–270.

55. Hamer M, Lavoie KL, Bacon SL. Taking up physical activity in later life and healthy ageing: the English longitudinal study of ageing. Br J Sports Med. 2014;48:239–243.

56. Schisterman EF, Cole SR, Platt RW. Overadjustment Bias and Unnecessary Adjustment in Epidemiologic Studies. Epidemiology. 2009;20:488–495.

57. Marconcin P, Werneck AO, Peralta M, Ihle A, Gouveia ÉR, Ferrari G, et al. The association between physical activity and mental health during the first year of the COVID-19 pandemic: a systematic review. BMC Public Health. 2022;22:209.

58. Hamer M, Stamatakis E, Steptoe A. Dose-response relationship between physical activity and mental health: the Scottish Health Survey. Br J Sports Med. 2009;43:1111–1114.

59. Pearce M, Garcia L, Abbas A, Strain T, Schuch FB, Golubic R, et al. Association Between Physical Activity and Risk of Depression: A Systematic Review and Meta-analysis. JAMA Psychiatry. 2022;79:550.

60. Schuch FB, Vancampfort D, Rosenbaum S, Richards J, Ward PB, Veronese N, et al. Exercise for depression in older adults: a meta-analysis of randomized controlled trials adjusting for publication bias. Braz J Psychiatry. 2016;38:247–254.

61. Jacobson NC, Newman MG. Anxiety and depression as bidirectional risk factors for one another: A meta-analysis of longitudinal studies. Psychol Bull. 2017;143:1155–1200.

62. Lamers F, van Oppen P, Comijs HC, Smit JH, Spinhoven P, van Balkom AJLM, et al. Comorbidity Patterns of Anxiety and Depressive Disorders in a Large Cohort Study: the Netherlands Study of Depression and Anxiety (NESDA). J Clin Psychiatry. 2011;72:341– 348.

63. Cho SH, Lim J-E, Lee J, Lee JS, Jeong H-G, Lee M-S, et al. Association between high-sensitivity C-reactive protein levels and depression: Moderation by age, sex, obesity, and aerobic physical activity. J Affect Disord. 2021;291:375–383.

64. Kruk J, Kotarska K, Aboul-Enein BH. Physical exercise and catecholamines response: benefits and health risk: possible mechanisms. Free Radic Res. 2020;54:105–125.

65. Beserra AHN, Kameda P, Deslandes AC, Schuch FB, Laks J, Moraes HS de. Can physical exercise modulate cortisol level in subjects with depression? A systematic review and meta-analysis. Trends Psychiatry Psychother. 2018;40:360–368.

66. Zhang J, Yao W, Hashimoto K. Brain-derived Neurotrophic Factor (BDNF)-TrkB Signaling in Inflammation-related Depression and Potential Therapeutic Targets. Curr Neuropharmacol. 2016;14:721–731.

67. Papathanassoglou EDE, Miltiadous P, Karanikola MN. May BDNF Be Implicated in the Exercise-Mediated Regulation of Inflammation? Critical Review and Synthesis of Evidence. Biol Res Nurs. 2015;17:521–539.

68. Byrne NJ, Soni S, Takahara S, Ferdaoussi M, Al Batran R, Darwesh AM, et al. Chronically Elevating Circulating Ketones Can Reduce Cardiac Inflammation and Blunt the Development of Heart Failure. Circ Heart Fail. 2020;13:e006573.

69. Chen L, Miao Z, Xu X. β-hydroxybutyrate alleviates depressive behaviors in mice possibly by increasing the histone3-lysine9-β-hydroxybutyrylation. Biochem Biophys Res Commun. 2017;490:117–122.

70. Kwak SE, Bae JH, Lee JH, Shin HE, Zhang D, Cho SC, et al. Effects of exercise-induced beta-hydroxybutyrate on muscle function and cognitive function. Physiol Rep. 2021;9.

71. Banks WA. The blood-brain barrier in neuroimmunology: Tales of separation and assimilation. Brain Behav Immun. 2015;44:1–8.

72. Eklund CM. Chapter 5 Proinflammatory cytokines in CRP baseline regulation. Adv. Clin. Chem., vol. 48, Elsevier; 2009. p. 111–136.

73. Rothaug M, Becker-Pauly C, Rose-John S. The role of interleukin-6 signaling in nervous tissue. Biochim Biophys Acta BBA - Mol Cell Res. 2016;1863:1218–1227.

74. Turkheimer FE, Althubaity N, Schubert J, Nettis MA, Cousins O, Dima D, et al. Increased serum peripheral C-reactive protein is associated with reduced brain barriers permeability of TSPO radioligands in healthy volunteers and depressed patients: implications for inflammation and depression. Brain Behav Immun. 2021;91:487–497.

75. Kelfve S, Kivi M, Johansson B, Lindwall M. Going web or staying paper? The use of web-surveys among older people. BMC Med Res Methodol. 2020;20:252.

76. Dyrstad SM, Hansen BH, Holme IM, Anderssen SA. Comparison of Self-reported versus Accelerometer-Measured Physical Activity. Med Sci Sports Exerc. 2014;46:99–106.

77. VanderWeele TJ. A Unification of Mediation and Interaction: A 4-Way Decomposition. Epidemiology. 2014;25:749–761.

