## Supplementary Materials for "Physical activity, low-grade inflammation, and psychological responses to the COVID-19 pandemic among older adults in England"

#### Supplement 1: Methods

##### Multiple imputation

MICE estimates the missing values under the missing-at-random assumption, meaning that the probability of missingness depends on observed variables [1]. Our final dataset had the highest proportion of missing data in the outcome variables (27.2%), whereas data on pre-pandemic physical activity levels were available for nearly all participants. Biomarkers were missing either due to unavailable or incomplete blood samples or bioassay errors. Details are outlined in the pre-registration protocol [2] and Supplementary Table 2. Issues during blood sample collection were recorded and accounted for in the imputation models. Bioassay errors are generally considered as independent of participant characteristics; therefore, this portion of data were missing completely at random and could be reliably imputed to increase power [3, 4]. Other variables were missing due to attrition, errors, or incomplete interviews.

The imputation models were specified on a variable-by-variable basis (fully conditional specification), ensuring each imputation model included all variables used in the corresponding substantive (analytical) model. Interaction terms between PA or PA changes and LGI were included using passive imputation. Other variables included issues during blood sample collection and previously identified drivers of attrition measured at baseline, namely housing tenure, occupational category, self-reported health, baseline working status, and self-rated memory. We also included two auxiliary variables (having a positive COVID-19 test and loneliness) from the first wave of the COVID-19 sub-study for variables with correlation r >0.1 to increase imputation precision.  Two sets of imputed datasets were constructed this way, each set using the same variables as part of the imputation models, differing only in the inclusion of pre-pandemic PA (used for models 1-3) or PA changes (used for models 4-5).

**Inference criteria**

The *s*-value can be interpreted as evidence against the null hypothesis expressed as the number of heads in a fair coin toss ($s = -\log_2 p$). To illustrate, $s = 4$ means that, assuming the true effect is null, observing the given test statistic or a more extreme test statistic would be as likely as obtaining four heads in a row in a fair coin toss. Unlike *p*-values, *s*-values scale more intuitively, with larger *s*-values providing more evidence against the null hypothesis [5].

95% confidence intervals for *OR*s were interpreted as compatibility intervals as described by Amrhein et al. [6]. Therefore, we considered effects included in a confidence interval as highly compatible with our data under a given statistical model. The confidence interval widths were used to assess the precision of *OR* point estimates [7].

Sensitivity power analyses were conducted in G*Power v3.1.9.6 [8]. *MDE* is the smallest effect that could be reliably detected with a given sample size and power [9]. Power was set to 0.8. *MDE*s were compared to the estimated effect sizes. If other metrics found little evidence for an effect, we assessed if effects smaller than *MDE*s would be theoretically meaningful. Required model parameters were obtained from the data and the imputed models. *MDE*s could only be obtained for the main effects, as the methods implemented in G*Power are not appropriate for logistic regression with interactions [10].

### Supplement 2: Sensitivity analyses

Firstly, the effect sizes and their confidence intervals from complete-case analyses were comparable to the pooled estimates from imputed datasets, suggesting a low risk of bias from imputing the data (Supplementary Table 6). However, the effect of pre-pandemic PA on anxiety was imprecise, with confidence interval not consistent in one direction of association. Secondly, additional adjustment for self-rated weight provided results similar to the main analyses (Supplementary Table 7). Thirdly, sensitivity analyses using continuous outcome scores also found a pattern consistent with the main analyses (Supplementary Table 8). Fourth, weighted analyses using Wave 9 measures of PA found slightly stronger associations between pre-pandemic PA and depression/anxiety (Supplementary Table 9). Fifth, the main results remained similar to those obtained from analyses that followed the pre-registration protocol (Supplementary Table 10). Lastly, we found comparable effects when recoding PA and its changes into binary variables indicating moderate-to-vigorous PA engagement at least once a week (Supplementary Table 11).

**Supplementary Fig. 1: Participant Flowchart**

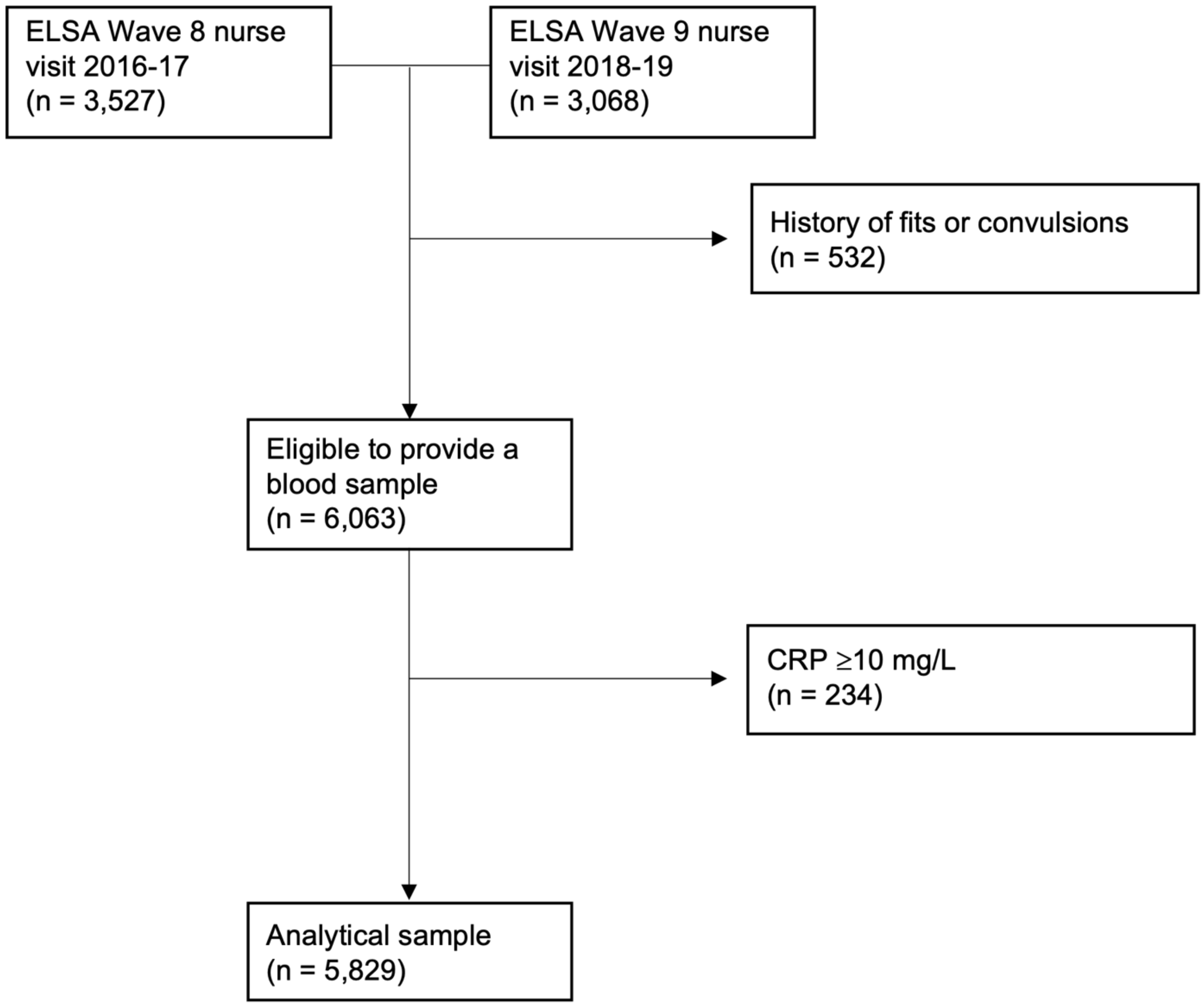


*Note.* The analytic sample included participants who (1) attended the nurse visit at Wave 8 or Wave 9 of ELSA, (2) did not have a history of blood fits or convulsions (thus were eligible to provide a blood sample), (3) had levels of CRP <10 mg/L.

**Supplementary Fig. 2: Measures Flowchart**

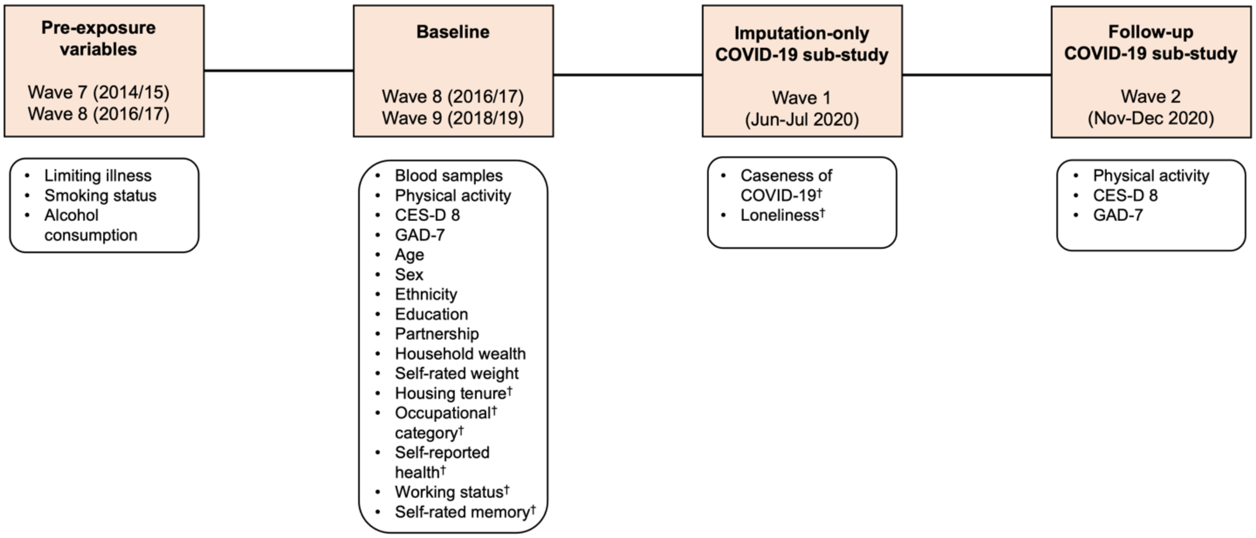


*Note.* Due to financial reasons, only some participants were offered the nurse visit at ELSA Wave 8, with the rest of the participants providing blood samples at Wave 9 instead. Consequently, baseline measures for each participant were extracted from either Wave 8 or Wave 9, as appropriate. CES-D 8 = Center for Epidemiologic Studies Depression scale; GAD-7: Generalised Anxiety Disorder scale.

† Used for imputation purposes only.

**Supplementary Fig. 3: Physical activity engagement before and during the COVID-19 pandemic**

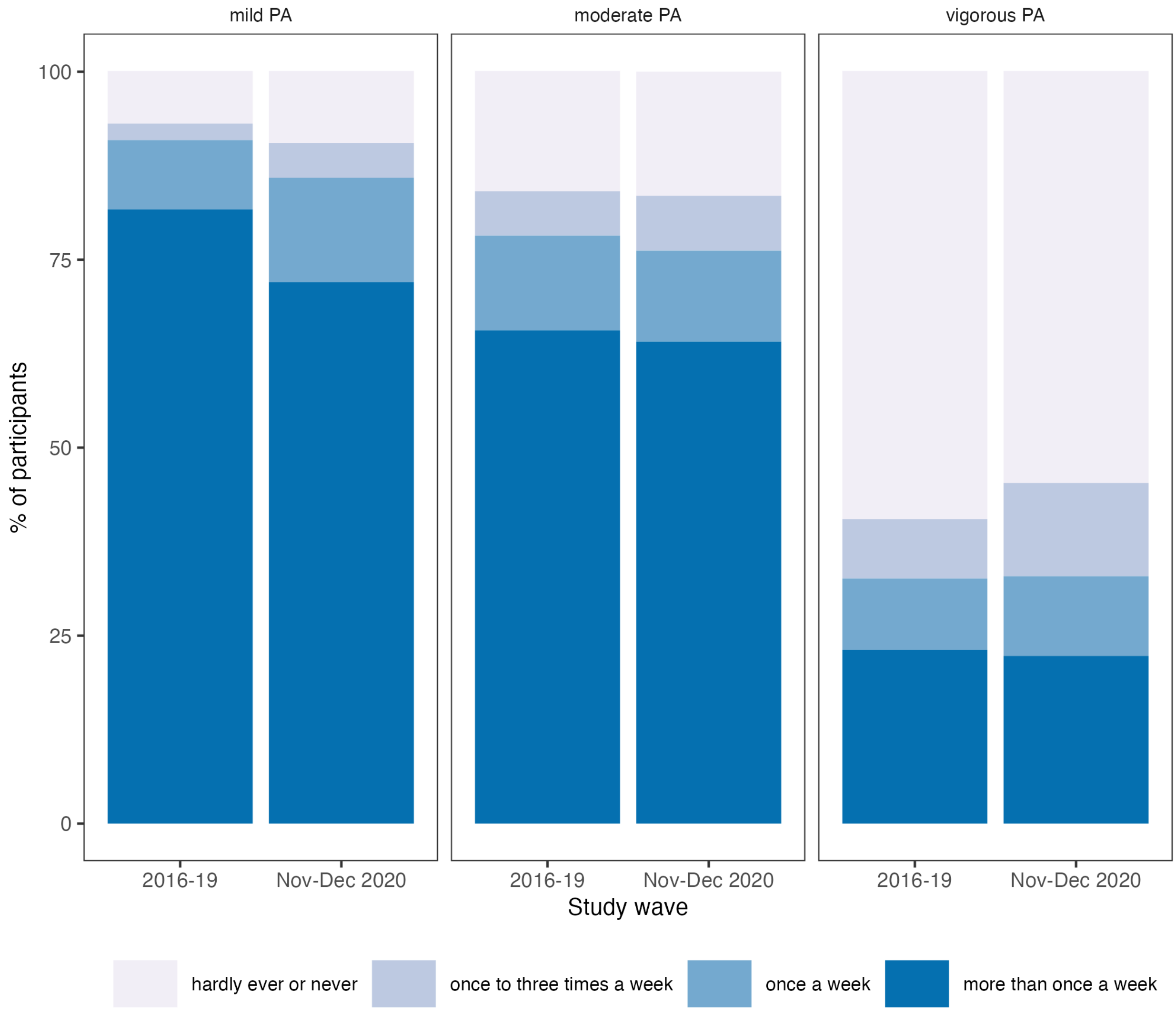


*Note*. The figure shows self-reported engagement in three intensities of physical activity: mild, moderate, and vigorous. Participants indicated how often they engaged in each of the three intensities, with response options including 'hardly ever, or never', 'once to three times a week', 'once a week', and 'more than once a week'. Pre-pandemic physical activity was obtained from Wave 8 (2016/17) and Wave 9 (2018/19) of ELSA depending on each participant, whereas pandemic data were acquired at Wave 2 of the COVID-19 sub-study (Nov-Dec 2020). PA = physical activity.

**Supplementary Table 1: Amendments to the pre-registration protocol**

| Item | Original | Revised | Reason for change |
|---|---|---|---|
| **Inclusion criteria** | 1. Attend W8 or W9 nurse visit.<br>2. Be eligible to provide a blood sample.<br>3. CRP ≤10 mg/L at baseline.<br>4. Attend W9 main interview.<br>5. Attend COVID-19 W1 interview.<br>6. Attend COVID-19 W2 interview. | 1. Attend W8 or W9 nurse visit.<br>2. Be eligible to provide a blood sample.<br>3. CRP ≤10 mg/L. | Running a completer analysis could lead to selection bias. We decided to extend our imputations to address attrition (see below). Therefore, participants were only required to attend the nurse visit at baseline. |
| **Imputations** | • Imputed missing data for completers.<br>• MICE used 20 imputed datasets and iterations. | • Imputed missing outcome values for those lost to attrition as well.<br>• Increased the number of imputations and iterations to 30.<br>• Included previously identified drivers of attrition in the imputation models (housing tenure, occupation, self-reported health, working status, self-rated memory) | The reason was to reduce selection bias due to attrition. Numbers of imputations and iterations were increased due to a larger proportion of missing data in the outcome variable (27.2%). |
| **Depression outcome** | • CES-D 8 (7-item version) | • CES-D 8 (full 8-item version) | During the time of pre-registration, our understanding was that the full CES-D 8 scale was only available for the main ELSA waves, whereas one item was accidentally dropped from the COVID-19 interviews. Upon closer inspection of the datasets, this was only true for COVID-19 W1. Given that the outcome of interest was measured at COVID-19 W2, we included the full CES-D 8 scale. |
| **Exposure timing** | • CRP at W8/W9<br>• PA at W9<br>• PA difference from W9 to W11 | • CRP at W8/W9<br>• PA at W8/W9<br>• PA difference from W8/W9 to W11 | The original baseline for PA was set at W9, reflecting PA levels immediately prior to the COVID-19 pandemic. However, this could introduce inconsistencies in the timing of confounders and exposures. CRP was assessed at W8 for some participants and at W9 for others. To maintain consistency across the models, we decided to define the baseline as W8/W9 for all variables, respectively. |
| **Change in exposure** | • Use change score ($PA_{W11}$ – $PA_{W9}$) as the exposure. | • Use $PA_{W11}$ as the exposure and derive the effects of change in PA using adjustment for $PA_{W8/W9}$ | Between the conception and finalisation of this project, change scores have been scrutinised in various contexts [11–13]. We have decided to follow the approach outlined by Katsoulis et al. [12], so that the change-in-exposure estimand has a clearer interpretation. |
| **Baseline confounders** | Baseline confounders at W9<br>• Depression (CES-D 8 / ONS)<br>• Age<br>• Partnership<br>• Wealth<br>• Limiting longstanding illness | Baseline confounders at W8/W9<br>• Depression (CES-D 8 / ONS)<br>• Age<br>• Partnership<br>• Wealth<br>+ Wave indicator | The baseline was set to either W8 or W9, depending on the participant. We added a wave indicator to account for the different baselines used for each participant. Adjusting for the presence of a limiting illness at baseline might lead to overadjustment bias, as it could be situated on the causal pathway between CRP, PA, and mental health. |
| **Covariates with unclear role (confounders or mediators)** | • Run sensitivity analyses adjusting for baseline (W9) smoking status, alcohol consumption, and self-rated weight.<br>• Adjust the main analyses for having a positive COVID-19 test result. | • Adjust the main analyses for smoking status, alcohol consumption, and limiting longstanding illness measured immediately prior to the exposure value (W7/W8).<br>• Run a sensitivity analysis adjusting for self-rated weight at baseline (W8/W9) | Adjusting for past levels of confounders prevents overadjustment bias, as past confounders cannot be retroactively affected by future exposure levels. However, W7 and W8 offered different adiposity measures. Therefore, we conducted a sensitivity analysis that adjusted for self-rated weight at baseline. |

| | | | |
|---|---|---|---|
| **Caseness of COVID-19 (used for imputations)** | • Indicated by self-reports of a positive COVID-19 test. | • Indicated by any of the following: a self-reported positive test for the coronavirus, staying in hospital due to coronavirus, or the reporting of at least two of the three core symptoms since the start of the pandemic (high temperature, a new continuous cough, loss of smell). | In the ELSA COVID-19 W1 dataset, only 23 participants reported testing positive for the coronavirus. Therefore, we derived a more comprehensive measure to indicate whether a person was likely to have experienced COVID-19. |
| **Plots** | • Results plotted as predicted probabilities with confounders set to their mean (continuous) or mode (categorical) values. | • Results plotted as average counterfactual predicted probabilities standardised to the distribution of confounders. | Setting confounders to their mean/mode values yields conditional predictions. These predictions may not represent population-level effects, limiting their generalisability to a hypothetical average person. |
| **Effect measures** | • Use logistic regression to estimate ORs, 95% CIs, and p-values.<br>• Compute *MDE*s for all models. | • Use logistic regression to estimate ORs, 95% CIs, and p-values.<br>• Use Poisson regression with a sandwich variance estimator to approximate RRs with 95% CIs.<br>• Compute *MDE*s for models with main effects. | The outcome was common (>10%), so *ORs* can overestimate the relative risk and may become difficult to interpret. For this reason, we assessed if *RRs* yielded similar results. Computing *MDE*s for interaction terms accurately would require more complex methods, such as power simulations. |
| **Sensitivity analyses** | • Rerun using the unimputed dataset,<br>• Rerun with binary coding of PA.<br>• Rerun with linear regression.<br>• Rerun with additional adjustment for self-rated weight, smoking, and alcohol consumption. | • Rerun using the unimputed dataset.<br>• Rerun with binary coding of PA.<br>• Rerun with linear regression.<br>• Rerun with additional adjustment for self-rated weight.<br>• Rerun analyses of PA using W9 measures from all participants who attended the COVID-19 W2, using longitudinal W9-COVID W2 weights.<br>• Rerun the main analyses exactly as pre-registered. | For amendments relating to the additional confounder adjustment, see the section on confounders. The additional sensitivity analysis using weights examined physical activity levels immediately before the pandemic and addressed selection bias (attending the nurse visit, population representativeness of the ELSA sample). |

**Supplementary Table 2: Overview of the covariates**

| Variable | Wave | Description | Coding |
| --- | --- | --- | --- |
| **Pre-pandemic mental health** | | | |
| Depression | W8/W9 | Center for Epidemiologic Studies Depression Scale (CES-D 8) | CES-D ≥4 |
| Anxiety | W8/W9 | ONS-4 anxiety item | Continuous |
| **Sociodemographic** | | | |
| Age at baseline | W8/W9 | - | Continuous (years) |
| Gender | W8/W9 | - | Female/Male |
| Ethnicity | W8/W9 | Self-reported ethnicity | White/Non-White |
| Education | W8/W9 | Self-reported highest achieved qualifications harmonized as part of ELSA project. | Less than upper secondary / Upper secondary & vocational training / Tertiary |
| Partnership | W8/W9 | - | Partnered (Married or Partnered) / Not partnered (Separated or Divorced or Widowed or Never Married) |
| Household wealth | W8/W9 | Index variable derived using over 20 components measured in ELSA, including investments and savings, housing, and debt. See Marmot et al. [14] for details. | Tertiles (obtained separately for W8 and W9) |
| **Health-related** | | | |
| Limiting longstanding illness | W7/W8 | Answering yes to both of the following:<br>1. *"Do you have any longstanding illness, disability or infirmity? By long-standing, I mean anything that has troubled you over a period of time."*<br>2. *"Does this illness or disability limit your activities in any way?"* | Yes/No |
| Smoking status | W7/W8 | Having reported smoking at all nowadays | Yes/No |
| Alcohol consumption | W7/W8 | How often a participant reported having a drink in the past 12 months ('not at all in the last 12 months', 'once or twice a year', 'once every couple of months', 'once or twice a month', 'once or twice a week', 'three or four days a week', 'five or six days a week', 'almost every day'). | Treated as continuous in the regression models (0-7), reported collapsed categories for descriptive purposes. |
| Self-rated weight (sensitivity analysis) | W8/W9 | Participants were asked *"Given your age and height, would you say that you are: about the right weight / too heavy / too light"* | About the right weight / Too heavy / Too light |
| **Imputation-only** | | | |
| Probable caseness of COVID-19 | COVID-19 W1 | Having experienced any of the following at any point since the start of the pandemic: a positive test for coronavirus, having to stay in hospital due to coronavirus, having developed at least two of the following symptoms: high fever, loss of smell, continuous new cough | Yes/No |
| Loneliness | COVID-19 W1 | Revised 20-item UCLA loneliness scale: | Continuous (sum score) |
| Housing tenure | W8/W9 | Participants were asked whether the current accommodation is owned, rented, or occupied. | Owning (Owning outright, Buying with a mortgage or loan, Paying rent & part of the mortgage) / Not owning (Renting, Living there rent free, Squatting) |
| Occupational category | W8/W9 | Participants were asked about their occupation. These categories were then collapsed using the NS-SEC 3 category classification as part of the ELSA project. | Managerial and Professional / Intermediate / Routine and Manual |
| Self-rated memory | W8/W9 | Participants were asked to rate their memory at the present time. | Excellent / Very good / Good / Fair / Poor |

| | | | |
|---|---|---|---|
| Self-rated health | W8/W9 | Participants were asked to rate their health at the present time. | Excellent / Very good / Good / Fair / Poor |
| Working status | W8/W9 | Self-reported working situation | Part of the labour force (employed, self-employed, unemployed or partly retired) / Not part of the labour force (retired, disabled, or looking after home or family) |
| Collapsing or poor veins | W8/W9 | Nurse-recorded problems during blood sample collection due to a participant having poor veins or collapsing. | Yes/No |
| Incomplete blood sample | W8/W9 | Nurse-recorded problems during blood sample collection due to the blood sample being incomplete. | Yes/No |
| Second attempt at taking blood needed | W8/W9 | Nurse-recorded problems during blood sample collection due to having to attempt blood sample collection for the second time. | Yes/No |

**Supplementary Table 3: Descriptive statistics for the unimputed and imputed datasets**

| | Complete dataset | | | Datasets imputed using pre-pandemic PA | | Datasets imputed using changes in PA | |
|---|---|---|---|---|---|---|---|
| **Characteristic** | **Missing (%)** | **% / Mean (*SD*)** | **Median (*IQR*)†** | **% / Mean (*SD*)** | **Median (*IQR*)** | **% / Mean (*SD*)** | **Median (*IQR*)** |
| **Sociodemographic characteristics** | | | | | | | |
| Sex: Female | None | 57.0 | - | 57.0 | - | 57.0 | - |
| Age (yrs) | None | 67.9 (9.9) | - | 67.9 (9.9) | - | 67.9 (9.9) | - |
| Ethnicity: White | <0.001 | 96.0 | - | 96.0 | - | 96.0 | - |
| Partnership: Partnered | <0.001 | 68.0 | - | 68.0 | - | 68.0 | - |
| Education | 8.8 | - | - | - | - | - | - |
| *Less than upper secondary* | - | 22.8 | - | 23.1 | - | 23.2 | - |
| *Upper secondary and vocational training* | - | 55.0 | - | 55.0 | - | 55.0 | - |
| *Tertiary* | - | 22.2 | - | 21.9 | - | 21.8 | - |
| Wealth tertiles | 6.3 | - | - | - | - | - | - |
| *First* | - | 33.3 | - | 33.6 | - | 33.5 | - |
| *Second* | - | 34.4 | - | 34.4 | - | 34.5 | - |
| *Third* | - | 32.3 | - | 32.0 | - | 32.1 | - |
| **Health-related factors** | | | | | | | |
| Limiting longstanding illness | 13.8 | 30.2 | - | 29.2 | - | 29.0 | - |
| Smoker | 13.8 | 9.3 | - | 11.0 | - | 10.8 | - |
| Alcohol consumption | 21.2 | | | | | | |
| *Three or more times a week* | - | 34.3 | - | 33.7 | - | 33.7 | - |
| *Once or twice a week* | - | 24.9 | - | 24.7 | - | 24.9 | - |
| *Less than once a week or not at all in a year* | - | 40.7 | - | 41.6 | - | 41.4 | - |
| **Exposures** | | | | | | | |
| Low-grade inflammation *(hsCRP ≥3 mg/L)* | 17.7 | 22.5 | | 23.9 | - | 23.8 | - |
| *hsCRP (mg/L)* | 17.7 | 2.0 (2.0) | 1,2 (2.1) | NI | - | NI | - |
| Physical activity (PA) | | | | | | | |
| *PA index (pre-pandemic)* | <0.001 | 10.1 (5.5) | - | 10.1 (5.5) | - | NI | - |
| *PA index (pandemic)* | 26.2 | 10.0 (5.4) | - | 9.5 (5.5) | - | NI | - |
| *PA index difference* | 26.2 | -0.8 (5.2) | - | NI | - | -0.8 (5.2) | - |
| **Mental health outcomes** | | | | | | | |
| Depressive symptoms (pre-pandemic) | | | | | | | |
| *Elevated (CES-D 8 ≥4)* | 6.6 | 11.4 | - | 11.5 | - | 11.5 | - |
| *CES-D 8* | 6.6 | 1.3 (1.8) | 1.0 (2.0) | NI | NI | NI | NI |
| Depressive symptoms (pandemic) | | | | | | | |
| *Elevated (CES-D 8 ≥4)* | 27.2 | 23.9 | - | 25.8 | - | 25.8 | - |
| *CES-D 8* | 27.2 | 2.1 (2.3) | 1.0 (3.0) | NI | NI | NI | NI |
| Anxiety symptoms | | | | | | | |
| *ONS-4 (pre-pandemic)* | 10.3 | 2.5 (2.6) | 2.0 (4.0) | 2.5 (2.6) | 2.0 (4.0) | 2.5 (2.6) | 2.0 (4.0) |
| *Elevated (pandemic, GAD-7 ≥10)* | 27.2 | 8.8 | - | 10.0 | - | 10.0 | - |
| *GAD-7 (pandemic)* | 27.2 | 3.2 (4.2) | 2.0 (5.0) | NI | NI | NI | NI |
| **Additional confounders** | | | | | | | |
| Self-rated weight | 0.5 | - | - | - | - | - | - |
| *Too light* | - | 3.2 | - | 3.8 | - | 3.8 | - |
| *About the right weight* | - | 44.4 | - | 44.0 | - | 44.0 | - |
| *Too heavy* | - | 52.4 | - | 52.2 | - | 52.2 | - |

*Note.* Descriptive statistics were obtained from complete-case and multiply imputed datasets. Two imputation models were used to estimate the missing data values depending on the substantive (analytical model). Model 1 included pre-pandemic physical activity, whilst Model 2 used changes in PA from before to during the pandemic. Both imputation models additionally included all other variables and interaction terms used in the main analytical models and auxiliary variables accounting for missingness. The descriptive statistics were averaged over 30 imputed datasets generated per imputation model. *IQR* = interquartile range; NI = not imputed; *SD* = standard deviation. † Listed for continuous variables with skewness or kurtosis ≥1.

**Supplementary Table 4: Characteristics of the full ELSA Wave 8 and Wave 9 samples and the analytical sample**

| | ELSA Wave 8 (N = 8,445) | | ELSA Wave 9 (N = 8,736) | | Analytical sample (N = 5,829) | |
|---|---|---|---|---|---|---|
| | **% / Mean (*SD*)** | **Median (*IQR*)†** | **% / Mean (*SD*)** | **Median (*IQR*)†** | **% / Mean (*SD*)** | **Median (*IQR*)†** |
| **Sociodemographic characteristics** | | | | | | |
| Sex: Female | 55.6 | - | 55.9 | - | 57.0 | - |
| Age (yrs) | 68.9 (9.6) | - | 67.8 (10.7) | - | 67.9 (9.9) | - |
| Ethnicity: White | 96.3 | - | 95.1 | - | 96.0 | - |
| Partnership: Partnered | 71.0 | - | 71.3 | - | 68.0 | - |
| Education | | | | | | |
| *Less than upper secondary* | 27.1 | - | 23.1 | - | 22.8 | - |
| *Upper secondary and vocational training* | 52.3 | - | 54.5 | - | 55.0 | - |
| *Tertiary* | 20.6 | - | 22.4 | - | 22.2 | - |
| Household wealth | 200 817 (711 065) | 50 216 (170 200) | 206 234 (518 068) | 53 000 (192 470) | 195 662 (729 549) | 50 500 (167 491) |
| **Health-related factors** | | | | | | |
| Limiting longstanding illness | 32.4 | - | 33.2 | - | 30.2 | - |
| Smoker | 10.6 | - | 9.3 | - | 9.3 | - |
| Alcohol consumption | | | | | | |
| *Three or more times a week* | 33.9 | - | 34.1 | - | 34.3 | - |
| *Once or twice a week* | 24.9 | - | 23.6 | - | 24.9 | - |
| *Less than once a week or not at all in a year* | 41.2 | - | 42.3 | - | 40.7 | - |
| **Mental health** | | | | | | |
| Depressive symptoms (pre-pandemic) | | | | | | |
| *Elevated (CES-D 8 ≥4)* | 12.7 | - | 12.3 | - | 11.4 | - |
| *CES-D 8* | 1.3 (1.8) | 1.0 (2.0) | 1.4 (1.8) | 1.0 (1.0) | 1.3 (1.8) | 1.0 (2.0) |
| Anxiety symptoms (pre-pandemic) | | | | | | |
| *ONS-4* | 2.6 (2.6) | 2.0 (4.0) | 2.4 (2.7) | 2.0 (4.0) | 2.5 (2.6) | 2.0 (4.0) |

*Note*. Descriptive statistics were obtained from complete-case datasets. The sample sizes refer to the overall sample.
† Listed for continuous variables with skewness or kurtosis >1.0

**Supplementary Table 5: Unadjusted and adjusted odds and risk ratios for the main models**

| Variable | $OR_{crude}$ [95% *CI*] | $OR_{adjusted}$ [95% *CI*] | $RR_{crude}$ [95% *CI*] | $RR_{adjusted}$ [95% *CI*] |
|---|---|---|---|---|
| **Depression** | | | | |
| Model 1 | | | | |
| *LGI* | 1.611 [1.354; 1.918] | 1.343 [1.100; 1.641] | 1.408 [1.248; 1.589] | 1.200 [1.066; 1.350] |
| Model 2 | | | | |
| *PA* | 0.932 [0.918; 0.945] | 0.964 [0.948; 0.981] | 0.949 [0.94; 0.959] | 0.977 [0.966; 0.988] |
| Model 3 | | | | |
| *LGI* | 1.531 [1.073; 2.183] | 1.367 [0.911; 2.052] | 1.279 [1.028; 1.591] | 1.147 [0.934; 1.408] |
| *PA* | 0.939 [0.923; 0.955] | 0.969 [0.950; 0.988] | 0.953 [0.942; 0.965] | 0.978 [0.966; 0.990] |
| *LGI * PA* | 0.989 [0.958; 1.022] | 0.993 [0.959; 1.029] | 0.998 [0.977; 1.020] | 1.002 [0.983; 1.023] |
| Model 4 | | | | |
| *PA change* | 0.946 [0.929; 0.963] | 0.983 [0.966; 1.000] | 0.992 [0.98; 1.003] | 0.960 [0.948; 0.973] |
| Model 5 | | | | |
| *LGI* | 1.455 [1.094; 1.936] | 1.355 [1.117; 1.645] | 1.421 [1.261; 1.600] | 1.232 [1.034;1.468] |
| *PA change* | 0.951 [0.932; 0.971] | 0.983 [0.963; 1.003] | 0.991 [0.977; 1.005] | 0.963 [0.948; 0.978] |
| *LGI * PA change* | 0.987 [0.958; 1.018] | 1.003 [0.972; 1.036] | 1.004 [0.984; 1.025] | 0.997 [0.977; 1.018] |
| **Anxiety** | | | | |
| Model 1 | | | | |
| *LGI* | 1.243 [0.945; 1.636] | 0.904 [0.669; 1.222] | 1.215 [0.952; 1.550] | 0.924 [0.733; 1.166] |
| Model 2 | | | | |
| *PA* | 0.932 [0.914; 0.950] | 0.976 [0.953; 1.000] | 0.939 [0.923; 0.955] | 0.982 [0.964; 1.002] |
| Model 3 | | | | |
| *LGI* | 1.416 [0.861; 2.329] | 1.148 [0.660; 1.995] | 1.334 [0.879; 2.023] | 1.096 [0.742; 1.620] |
| *PA* | 0.941 [0.921; 0.962] | 0.984 [0.958; 1.010] | 0.947 [0.929; 0.966] | 0.988 [0.967; 1.009] |
| *LGI * PA* | 0.961 [0.912; 1.013] | 0.964 [0.911; 1.020] | 0.967 [0.924; 1.012] | 0.973 [0.932; 1.014] |
| Model 4 | | | | |
| *PA change* | 0.943 [0.919; 0.968] | 0.964 [0.937; 0.992] | 0.976 [0.953; 1.000] | 0.949 [0.927; 0.971] |
| Model 5 | | | | |
| *LGI* | 1.165 [0.751; 1.807] | 0.933 [0.699; 1.246] | 1.253 [1.001; 1.570] | 1.128 [0.784; 1.622] |
| *PA change* | 0.948 [0.922; 0.976] | 0.943 [0.934; 0.991] | 0.972 [0.948; 0.997] | 0.953 [0.929; 0.978] |
| *LGI * PA change* | 0.979 [0.931; 1.029] | 1.009 [0.961; 1.060] | 1.015 [0.976; 1.056] | 0.983 [0.941; 1.026] |

*Note.* The models show associations between clinically significant depressive or anxiety symptoms during the pandemic (November-December 2020) and pre-pandemic low-grade inflammation (2016/17 or 2018/2019; Model 1), pre-pandemic physical activity engagement (2016/17 or 2018/2019; Model 2), changes in physical activity from before to during the pandemic (Model 4), and interactions between inflammation and physical activity as indicated (Model 3 and 5). Odds ratios, standard errors, and p-values were obtained from logistic regression models. Risk ratios were approximated using Poisson regression with a sandwich variance estimator. Adjusted models accounted for pre-pandemic mental health (depression or anxiety depending on the outcome), sex, age, ethnicity, education, partnership status, household wealth, having a limiting longstanding illness, smoking status, and alcohol consumption. Effects of PA changes were estimated using coefficients of pandemic PA adjusted for pre-pandemic PA. *CI* = confidence interval; *LGI* = low-grade inflammation; *OR* = odds ratio; *PA* = physical activity.

**Supplementary Table 6: Adjusted logistic regression models using the unimputed dataset (sensitivity analysis)**

| Variable | *OR [95% CI]* | *SE* | *p* | *s* | *RR [95% CI]* |
|---|---|---|---|---|---|
| **Outcome: Depression** | | | | | |
| Model 1 (N = 2 636) | | | | | |
| *LGI* | 1.400 [1.110; 1.760] | 0.12 | 0.004 | 7.97 | 1.260 [1.090; 1.470] |
| Model 2 (N = 3 098) | | | | | |
| *PA* | 0.968 [0.949; 0.987] | 0.01 | 0.001 | 10.1 | 0.978 [0.965; 0.991] |
| Model 3 (N = 2 636) | | | | | |
| *LGI* | 1.690 [1.050; 2.720] | 0.24 | 0.030 | 5.05 | 1.320 [1.010; 1.720] |
| *PA* | 0.976 [0.953; 1.000] | 0.01 | 0.046 | 4.45 | 0.982 [0.965; 0.998] |
| *LGI * PA* | 0.976 [0.932; 1.020] | 0.02 | 0.288 | 1.79 | 0.992 [0.964; 1.020] |
| Model 4 (N = 3 097) | | | | | |
| *PA change* | 0.949 [0.929; 0.969] | 0.01 | <0.001 | 20.6 | 0.964 [0.950; 0.979] |
| Model 5 (N = 2 636) | | | | | |
| *LGI* | 1.810 [1.170; 2.810] | 0.22 | 0.008 | 6.99 | 1.380 [1.080; 1.750] |
| *PA change* | 0.957 [0.933; 0.982] | 0.01 | 0.001 | 10.5 | 0.968 [0.950; 0.986] |
| *LGI * PA change* | 0.959 [0.915; 1.000] | 0.02 | 0.082 | 3.61 | 0.980 [0.952; 1.010] |
| **Outcome: Anxiety** | | | | | |
| Model 1 (N = 2 513) | | | | | |
| *LGI* | 0.979 [0.669; 1.410] | 0.19 | 0.912 | 0.13 | 0.995 [0.735; 1.350] |
| Model 2 (N = 2 952) | | | | | |
| *PA* | 0.988 [0.958; 1.020] | 0.02 | 0.423 | 1.24 | 0.991 [0.965; 1.020] |
| Model 3 (N = 2 513) | | | | | |
| *LGI* | 1.250 [0.600; 2.580] | 0.37 | 0.542 | 0.88 | 1.270 [0.714; 2.270] |
| *PA* | 0.996 [0.958; 1.040] | 0.02 | 0.856 | 0.23 | 0.999 [0.967; 1.030] |
| *LGI * PA* | 0.970 [0.901; 1.040] | 0.04 | 0.416 | 1.27 | 0.970 [0.913; 1.030] |
| Model 4 (N = 2 951) | | | | | |
| *PA change* | 0.951 [0.921; 0.982] | 0.02 | 0.002 | 8.98 | 0.959 [0.932; 0.986] |
| Model 5 (N = 2 513) | | | | | |
| *LGI* | 1.540 [0.788; 2.960] | 0.34 | 0.202 | 2.31 | 1.390 [0.865; 2.230] |
| *PA change* | 0.975 [0.936; 1.010] | 0.02 | 0.210 | 2.25 | 0.979 [0.946; 1.010] |
| *LGI * PA change* | 0.935 [0.865; 1.010] | 0.04 | 0.087 | 3.52 | 0.949 [0.895; 1.000] |

*Note.* The models show associations between clinically significant depressive or anxiety symptoms during the pandemic (November-December 2020) and pre-pandemic low-grade inflammation (2016/17 or 2018/2019; Model 1), pre-pandemic physical activity engagement (2016/17 or 2018/2019; Model 2), changes in physical activity from before to during the pandemic (Model 4), and interactions between inflammation and physical activity as indicated (Model 3 and 5). Odds ratios, standard errors, and p-values were obtained from logistic regression models. Risk ratios were approximated using Poisson regression with a sandwich variance estimator. The models were adjusted for pre-pandemic mental health (depression or anxiety depending on the outcome), sex, age, ethnicity, education, partnership status, household wealth, having a limiting longstanding illness, smoking status, and alcohol consumption. Effects of PA changes were estimated using coefficients of pandemic PA adjusted for pre-pandemic PA and the outlined confounders. *CI* = confidence interval; *LGI* = low-grade inflammation; *OR* = adjusted odds ratio; *PA* = physical activity; RR = risk ratio, *s* = Shannon information value (surprisal value); *SE* = standard error.

**Supplementary Table 7: Results of adjusted logistic regression models with additional adjustment for self-rated weight (sensitivity analysis)**

| Variable | *OR [95% CI]* | *SE* | *p* | *s* | *RR [95% CI]* |
|---|---|---|---|---|---|
| **Outcome: Depression** | | | | | |
| Model 1 | | | | | |
| *LGI* | 1.262 [1.026; 1.552] | 0.10 | 0.028 | 5.17 | 1.150 [1.017; 1.301] |
| Model 2 | | | | | |
| *PA* | 0.967 [0.951; 0.984] | 0.01 | <0.001 | 12.72 | 0.978 [0.968; 0.990] |
| Model 3 | | | | | |
| *LGI* | 1.313 [0.868; 1.987] | 0.21 | 0.195 | 2.36 | 1.114 [0.901; 1.377] |
| *PA* | 0.971 [0.952; 0.990] | 0.01 | 0.004 | 8.15 | 0.979 [0.967; 0.992] |
| *LGI * PA* | 0.991 [0.957; 1.027] | 0.02 | 0.632 | 0.66 | 1.001 [0.981; 1.022] |
| Model 4 | | | | | |
| *PA change* | 0.957 [0.939; 0.976] | 0.01 | <0.001 | 16.11 | 0.972 [0.960; 0.985] |
| Model 5 | | | | | |
| *LGI* | 1.271 [0.911; 1.773] | 0.17 | 0.157 | 2.68 | 1.103 [0.925; 1.315] |
| *PA change* | 0.961 [0.940; 0.982] | 0.01 | 0.001 | 10.92 | 0.973 [0.959; 0.987] |
| *LGI * PA change* | 0.990 [0.958; 1.024] | 0.02 | 0.575 | 0.80 | 1.000 [0.980; 1.020] |
| **Outcome: Anxiety** | | | | | |
| Model 1 | | | | | |
| *LGI* | 0.839 [0.614; 1.148] | 0.16 | 0.271 | 1.89 | 0.871 [0.686; 1.107] |
| Model 2 | | | | | |
| *PA* | 0.978 [0.955; 1.002] | 0.01 | 0.072 | 3.79 | 0.983 [0.964; 1.003] |
| Model 3 | | | | | |
| *LGI* | 1.091 [0.618; 1.923] | 0.29 | 0.762 | 0.39 | 1.046 [0.703; 1.557] |
| *PA* | 0.986 [0.960; 1.012] | 0.01 | 0.290 | 1.80 | 0.989 [0.968; 1.010] |
| *LGI * PA* | 0.961 [0.908; 1.018] | 0.03 | 0.170 | 2.53 | 0.971 [0.931; 1.013] |
| Model 4 | | | | | |
| *PA change* | 0.956 [0.929; 0.984] | 0.02 | 0.002 | 8.66 | 0.966 [0.943; 0.988] |
| Model 5 | | | | | |
| *LGI* | 0.956 [0.573; 1.596] | 0.26 | 0.863 | 0.21 | 0.940 [0.651; 1.359] |
| *PA change* | 0.961 [0.931; 0.991] | 0.02 | 0.013 | 6.28 | 0.968 [0.944; 0.993] |
| *LGI * PA change* | 0.976 [0.927; 1.029] | 0.03 | 0.370 | 1.43 | 0.984 [0.947; 1.024] |

*Note.* The models show associations between clinically significant depressive or anxiety symptoms during the pandemic (November-December 2020) and pre-pandemic low-grade inflammation (2016/17 or 2018/2019; Model 1), pre-pandemic physical activity engagement (2016/17 or 2018/2019; Model 2), changes in physical activity from before to during the pandemic (Model 4), and interactions between inflammation and physical activity as indicated (Model 3 and 5). The results were pooled from 30 imputed datasets (sample N = 5,829). Odds ratios, standard errors, and p-values were obtained from logistic regression models. Risk ratios were approximated using modified Poisson regression with a sandwich variance estimator. The models were adjusted for pre-pandemic mental health (depression or anxiety depending on the outcome), sex, age, ethnicity, education, partnership status, household wealth, having a limiting longstanding illness, smoking status, alcohol consumption, and self-rated weight. Effects of PA changes were estimated using coefficients of pandemic PA adjusted for pre-pandemic PA and the outlined confounders. *CI* = confidence interval; *LGI* = low-grade inflammation; *OR* = adjusted odds ratio; *PA* = physical activity; *RR* = risk ratio, *s* = Shannon information value (surprisal value); *SE* = standard error.

**Supplementary Table 8: Results of adjusted linear regression models with sandwich variance estimator (sensitivity analysis)**

| Variable | *b* | 95% *CI* | *SE* | *p* | *s* |
|---|---|---|---|---|---|
| **Outcome: CES-D 7** | | | | | |
| Model 1 | | | | | |
| *LGI* | 0.231 | 0.049; 0.413 | 0.09 | 0.013 | 6.23 |
| Model 2 | | | | | |
| *PA* | -0.022 | -0.036; -0.009 | 0.01 | 0.002 | 9.23 |
| Model 3 | | | | | |
| *LGI* | 0.283 | -0.094; 0.660 | 0.19 | 0.140 | 2.84 |
| *PA* | -0.019 | -0.034; -0.003 | 0.01 | 0.019 | 5.75 |
| *LGI * PA* | -0.008 | -0.039; 0.022 | 0.02 | 0.591 | 0.76 |
| Model 4 | | | | | |
| *PA change* | -0.044 | -0.060; -0.028 | 0.01 | <0.001 | 22.18 |
| Model 5 | | | | | |
| *LGI* | 0.274 | -0.034; 0.581 | 0.16 | 0.081 | 3.63 |
| *PA change* | -0.040 | -0.056; -0.023 | 0.01 | <0.001 | 18.02 |
| *LGI * PA change* | -0.013 | -0.040; 0.015 | 0.01 | 0.372 | 1.43 |
| **Outcome: GAD-7** | | | | | |
| Model 1 | | | | | |
| *LGI* | -0.228 | -0.546; 0.09 | 0.16 | 0.159 | 2.65 |
| Model 2 | | | | | |
| *PA* | -0.03 | -0.055; -0.004 | 0.01 | 0.022 | 5.52 |
| Model 3 | | | | | |
| *LGI* | -0.113 | -0.821; 0.595 | 0.36 | 0.753 | 0.41 |
| *PA* | -0.04 | -0.07; -0.01 | 0.01 | 0.004 | 7.85 |
| *LGI * PA* | -0.017 | -0.076; 0.042 | 0.03 | 0.570 | 0.81 |
| Model 4 | | | | | |
| *PA change* | -0.046 | -0.078; -0.014 | 0.02 | 0.005 | 7.68 |
| Model 5 | | | | | |
| *LGI* | -0.070 | -0.735; 0.596 | 0.34 | 0.836 | 2.56 |
| *PA change* | -0.042 | -0.076; -0.007 | 0.02 | 0.018 | 5.81 |
| *LGI * PA change* | -0.029 | -0.088; 0.030 | 0.03 | 0.329 | 1.60 |

*Note.* The models show associations between depressive or anxiety symptoms during the pandemic (November-December 2020) and pre-pandemic low-grade inflammation (2016/17 or 2018/2019; Model 1), pre-pandemic physical activity engagement (2016/17 or 2018/2019; Model 2), changes in physical activity from before to during the pandemic (Model 4), and interactions between inflammation and physical activity as indicated (Model 3 and 5). The results were pooled from 30 imputed datasets (sample N = 5,829). The models were adjusted for pre-pandemic mental health (depression or anxiety depending on the outcome), sex, age, ethnicity, education, partnership status, household wealth, having a limiting longstanding illness, smoking status, and alcohol consumption. Effects of PA changes were estimated using coefficients of pandemic PA adjusted for pre-pandemic PA and the outlined confounders. *b* = regression coefficient (unstandardised); CI = confidence interval; *LGI* = low-grade inflammation; *PA* = physical activity; *s* = Shannon information value (surprisal value); *SE* = standard error.

**Supplementary Table 9: Results of weighted adjusted logistic regression models using W9 PA measures (sensitivity analysis)**

| Variable | *OR [95% CI]* | *SE* | *p* | *s* | *RR [95% CI]* |
|---|---|---|---|---|---|
| **Outcome: Depression** | | | | | |
| Model 2 | | | | | |
| *PA* | 0.954 [0.937; 0.972] | 0.01 | <0.001 | 19.95 | 0.973 [0.961; 0.984] |
| Model 4 | | | | | |
| *PA change* | 0.959 [0.940; 0.978] | 0.01 | <0.001 | 15.14 | 0.975 [0.963; 0.987] |
| **Outcome: Anxiety** | | | | | |
| Model 2 | | | | | |
| *PA* | 0.962 [0.934; 0.991] | 0.02 | 0.010 | 6.61 | 0.973 [0.951; 0.996] |
| Model 4 | | | | | |
| *PA change* | 0.959 [0.931; 0.988] | 0.02 | 0.005 | 7.60 | 0.971 [0.948; 0.994] |

*Note.* The models show associations between clinically significant depressive or anxiety symptoms during the pandemic (November-December 2020) and pre-pandemic physical activity engagement (2018/2019; Model 2) and changes in physical activity from before to during the pandemic (Model 4). The models were weighted to the population of interest at Wave 9 using longitudinal W9–COVID-19 W2 survey weights and the results were pooled from 30 imputed datasets (sample N = 5,378). Odds ratios, standard errors, and p-values were obtained from logistic regression models. Risk ratios were approximated using modified Poisson regression with a sandwich variance estimator. The models were adjusted for pre-pandemic mental health (depression or anxiety depending on the outcome), sex, age, ethnicity, education, partnership status, household wealth, having a limiting longstanding illness, smoking status, and alcohol consumption. Effects of PA changes were estimated using coefficients of pandemic PA adjusted for pre-pandemic PA and the outlined confounders. *CI* = confidence interval; *OR* = adjusted odds ratio; *PA* = physical activity; *RR* = risk ratio, *s* = Shannon information value (surprisal value); *SE* = standard error.

**Supplementary Table 10: Results of the pre-registered main adjusted logistic regression models (sensitivity analysis)**

| Variable | *OR* | 95% *CI* | *SE* | *p* | *s* | *MDE* |
|---|---|---|---|---|---|---|
| **Outcome: Depression** | | | | | | |
| Model 1 | | | | | | |
| *LGI* | 1.55 | 1.28; 1.89 | 0.10 | <0.001 | 16.41 | 1.32 |
| Model 2 | | | | | | |
| *PA* | 0.96 | 0.94; 0.98 | 0.01 | <0.001 | 19.78 | 0.98 |
| Model 3 | | | | | | |
| *LGI* | 1.77 | 1.20; 2.59 | 0.20 | 0.004 | 7.98 | |
| *PA* | 0.97 | 0.95; 0.99 | 0.01 | 0.002 | 9.52 | |
| *LGI * PA* | 0.98 | 0.94; 1.02 | 0.02 | 0.297 | 1.75 | 0.90 |
| Model 4 | | | | | | |
| *PA change* | 0.98 | 0.96; 0.99 | 0.01 | 0.005 | 7.56 | 0.98 |
| Model 5 | | | | | | |
| *LGI* | 1.56 | 1.28; 1.91 | 0.10 | <0.001 | 16.00 | |
| *PA change* | 0.98 | 0.96; 0.997 | 0.01 | 0.025 | 5.32 | |
| *LGI * PA change* | 1.00 | 0.97; 1.04 | 0.02 | 0.930 | 0.11 | 0.95 |
| **Outcome: Anxiety** | | | | | | |
| Model 1 | | | | | | |
| *LGI* | 0.96 | 0.70; 1.31 | 0.16 | 0.781 | 0.36 | 1.39 |
| Model 2 | | | | | | |
| *PA* | 0.95 | 0.93; 0.97 | 0.01 | <0.001 | 14.15 | 0.96 |
| Model 3 | | | | | | |
| *LGI* | 1.23 | 0.70; 2.15 | 0.29 | 0.480 | 1.06 | |
| *PA* | 0.96 | 0.93; 0.99 | 0.02 | 0.004 | 7.98 | |
| *LGI * PA* | 0.96 | 0.91; 1.02 | 0.03 | 0.188 | 2.41 | 0.94 |
| Model 4 | | | | | | |
| *PA change* | 0.99 | 0.97; 1.01 | 0.01 | 0.291 | 1.78 | 0.97 |
| Model 5 | | | | | | |
| *LGI* | 0.97 | 0.72; 1.30 | 0.15 | 0.838 | 0.26 | |
| *PA change* | 0.99 | 0.96; 1.02 | 0.02 | 0.379 | 1.40 | |
| *LGI * PA change* | 1.00 | 0.95; 1.07 | 0.03 | 0.969 | 0.05 | 0.93 |

*Note.* The models show associations between clinically significant depressive or anxiety symptoms during the pandemic (November-December 2020) and pre-pandemic low-grade inflammation (2016/17 or 2018/2019; Model 1), pre-pandemic physical activity engagement (2018/2019; Model 2), changes in physical activity from before to during the pandemic (Model 4), and interactions between inflammation and physical activity as indicated (Model 3 and 5). The results were pooled from 20 imputed datasets (sample N = 4,148). The models were adjusted for pre-pandemic mental health (depression or anxiety depending on the outcome), sex, age, ethnicity, education, partnership status, household wealth, having a limiting longstanding illness, and previous COVID-19 infection. *CI* = confidence interval; *LGI* = low-grade inflammation; *MDE* = minimum detectable effect; *OR* = adjusted odds ratio; *PA* = physical activity; *s* = Shannon information value (surprisal value); *SE* = standard error.

**Supplementary Table 11: Adjusted logistic regression models with binary coding of physical activity (sensitivity analysis)**

| Variable | Depression | | | | | Anxiety | | | | |
|---|---|---|---|---|---|---|---|---|---|---|
| | ***OR* [95% *CI*]** | ***RR* [95% *CI*]** | ***SE*** | ***p*** | ***s*** | ***OR* [95% *CI*]** | ***RR* [95% *CI*]** | ***SE*** | ***p*** | ***s*** |
| Model 2 | | | | | | | | | | |
| $PA_{mod/vig}$ | 0.762 [0.612; 0.948] | 0.859 [0.756; 0.975] | 0.11 | 0.015 | 6.03 | 0.803 [0.582; 1.109] | 0.864 [0.671; 1.112] | 0.16 | 0.181 | 2.47 |
| Model 3 | | | | | | | | | | |
| *LGI* | 1.389 [0.984; 1.963] | 1.175 [0.991; 1.394] | 0.18 | 0.062 | 4.02 | 1.093 [0.638; 1.872] | 1.076 [0.744; 1.557] | 0.27 | 0.743 | 0.43 |
| $PA_{mod/vig}$ | 0.809 [0.630; 1.039] | 0.876 [0.755; 1.016] | 0.13 | 0.096 | 3.38 | 0.878 [0.591; 1.304] | 0.931 [0.692; 1.252] | 0.20 | 0.515 | 0.96 |
| $LGI * PA_{mod/vig}$ | 0.897 [0.601; 1.338] | 0.990 [0.798; 1.227] | 0.20 | 0.592 | 0.76 | 0.744 [0.401; 1.383] | 0.785 [0.501; 1.230] | 0.31 | 0.347 | 1.53 |
| Model 4 | | | | | | | | | | |
| *Ref:* $\Delta PA_{high\text{-}high}$ | | | | | | | | | | |
| $\Delta PA_{low\text{-}high}$ | 1.092 [0.814; 1.465] | 1.079 [0.901; 1.294] | 0.15 | 0.556 | 0.85 | 1.139 [0.728; 1.784] | 1.111 [0.779; 1.586] | 0.23 | 0.566 | 0.82 |
| $\Delta PA_{high\text{-}low}$ | 1.681 [1.358; 2.080] | 1.393 [1.222; 1.588] | 0.11 | <0.001 | 18.61 | 1.543 [1.156; 2.060] | 1.422 [1.130; 1.790] | 0.15 | 0.003 | 8.22 |
| $\Delta PA_{low\text{-}low}$ | 1.994 [1.537; 2.586] | 1.464 [1.275; 1.681] | 0.13 | <0.001 | 21.42 | 1.768 [1.220; 2.561] | 1.484 [1.126; 1.956] | 0.19 | 0.003 | 8.48 |
| Model 5 | | | | | | | | | | |
| *Ref:* $\Delta PA_{high\text{-}high}$ | | | | | | | | | | |
| $\Delta PA_{low\text{-}high}$ | 1.001 [0.703; 1.424] | 1.025 [0.817; 1.287] | 0.18 | 0.996 | 0.01 | 0.925 [0.518; 1.651] | 0.937 [0.591; 1.486] | 0.29 | 0.791 | 0.34 |
| $\Delta PA_{high\text{-}low}$ | 1.623 [1.264; 2.085] | 1.373 [1.175; 1.606] | 0.13 | <0.001 | 12.56 | 1.572 [1.099; 2.248] | 1.451 [1.097; 1.919] | 0.18 | 0.013 | 6.21 |
| $\Delta PA_{low\text{-}low}$ | 1.830 [1.346; 2.488] | 1.432 [1.214; 1.690] | 0.16 | <0.001 | 12.93 | 1.821 [1.181; 2.806] | 1.489 [1.093; 2.030] | 0.22 | 0.007 | 7.19 |
| *LGI* | 1.205 [0.937; 1.550] | 1.145 [0.965; 1.359] | 0.13 | 0.146 | 2.77 | 0.741 [0.497; 1.105] | 0.788 [0.562; 1.104] | 0.20 | 0.141 | 2.83 |
| $LGI * \Delta PA_{low\text{-}high}$ | 1.242 [0.692; 2.229] | 1.116 [0.791; 1.575] | 0.30 | 0.467 | 1.10 | 1.971 [0.770; 5.043] | 1.730 [0.828; 3.615] | 0.48 | 0.156 | 2.68 |
| $LGI * \Delta PA_{high\text{-}low}$ | 1.073 [0.672; 1.714] | 1.012 [0.774; 1.323] | 0.24 | 0.767 | 0.38 | 0.996 [0.435; 2.278] | 0.980 [0.511; 1.880] | 0.42 | 0.992 | 0.01 |
| $LGI * \Delta PA_{low\text{-}low}$ | 1.163 [0.707; 1.915] | 1.004 [0.780; 1.293] | 0.25 | 0.551 | 0.86 | 1.060 [0.520; 2.161] | 1.091 [0.647; 1.840] | 0.36 | 0.871 | 0.20 |

The models show associations between clinically significant depressive or anxiety symptoms during the pandemic (November-December 2020) and pre-pandemic low-grade inflammation (2016/17 or 2018/2019; Model 1), pre-pandemic physical activity engagement (2016/17 or 2018/2019; Model 2), changes in physical activity from before to during the pandemic (Model 4), and interactions between inflammation and physical activity as indicated (Model 3 and 5). The results were pooled from 30 imputed datasets (sample N = 5,829). Odds ratios, standard errors, and p-values were obtained from logistic regression models. Risk ratios were approximated using modified Poisson regression with a sandwich variance estimator. Δ = change (from pre-pandemic to pandemic levels), *CI* = confidence interval; *LGI* = low-grade inflammation; *OR* = adjusted odds ratio; *PA* = physical activity; *RR* = risk ratio; *s* = Shannon information value (surprisal value); *SE* = standard error.

**References for Supplementary Materials**


1. Sterne JAC, White IR, Carlin JB, Spratt M, Royston P, Kenward MG, et al. Multiple imputation for missing data in epidemiological and clinical research: potential and pitfalls. BMJ. 2009;338:b2393–b2393.
2. Danka MN, Iob E. Physical activity, low-grade inflammation, and psychological responses to the COVID-19 pandemic among older adults in England. 2022. 1 November 2022. https://doi.org/10.17605/OSF.IO/XJFYZ.
3. Austin PC, White IR, Lee DS, van Buuren S. Missing Data in Clinical Research: A Tutorial on Multiple Imputation. Can J Cardiol. 2021;37:1322–1331.
4. Kontopantelis E, White IR, Sperrin M, Buchan I. Outcome-sensitive multiple imputation: a simulation study. BMC Med Res Methodol. 2017;17:2.
5. Greenland S. Valid *P* -Values Behave Exactly as They Should: Some Misleading Criticisms of *P* -Values and Their Resolution With *S* -Values. Am Stat. 2019;73:106–114.
6. Amrhein V, Trafimow D, Greenland S. Inferential Statistics as Descriptive Statistics: There Is No Replication Crisis if We Don’t Expect Replication. Am Stat. 2019;73:262–270.
7. Anderson AA. Assessing Statistical Results: Magnitude, Precision, and Model Uncertainty. Am Stat. 2019;73:118–121.
8. Faul F, Erdfelder E, Buchner A, Lang A-G. Statistical power analyses using G*Power 3.1: Tests for correlation and regression analyses. Behav Res Methods. 2009;41:1149–1160.
9. Lakens D. Sample Size Justification. Collabra Psychol. 2022;8:33267.
10. Demidenko E. Sample size and optimal design for logistic regression with binary interaction. Stat Med. 2008;27:36–46.
11. Tennant PWG, Arnold KF, Ellison GTH, Gilthorpe MS. Analyses of ‘change scores’ do not estimate causal effects in observational data. Int J Epidemiol. 2022;51:1604–1615.
12. Katsoulis M, Lai AG, Kipourou DK, Gomes M, Banerjee A, Denaxas S, et al. On the estimation of the effect of weight change on a health outcome using observational data, by utilising the target trial emulation framework. Int J Obes. 2023;47:1309–1317.
13. Tennant PWG, Tomova GD, Murray EJ, Arnold KF, Fox MP, Gilthorpe MS. Lord’s ‘paradox’ explained: the 50-year warning on the use of ‘change scores’ in observational data. 2023.
14. Marmot M, Banks J, Blundell R, Lessof C. Health, wealth and lifestyles of the older population in England: The 2002 English Longitudinal Study of Aging. London: The Institute for Fiscal Studies; 2003.
